# Association between living with children and outcomes from COVID-19: an OpenSAFELY cohort study of 12 million adults in England

**DOI:** 10.1101/2020.11.01.20222315

**Authors:** Harriet Forbes, Caroline E Morton, Seb Bacon, Helen I McDonald, Caroline Minassian, Jeremy P Brown, Christopher T Rentsch, Rohini Mathur, Anna Schultze, Nicholas J DeVito, Brian MacKenna, William J Hulme, Richard Croker, Alex J Walker, Elizabeth J Williamson, Chris Bates, Amir Mehrkar, Helen J Curtis, David Evans, Kevin Wing, Peter Inglesby, Henry Drysdale, Angel YS Wong, Jonathan Cockburn, Robert McManus, John Parry, Frank Hester, Sam Harper, Ian J Douglas, Liam Smeeth, Stephen JW Evans, Krishnan Bhaskaran, Rosalind M Eggo, Ben Goldacre, Laurie A Tomlinson

## Abstract

**Background:** Close contact with children may provide cross-reactive immunity to SARs-CoV-2 due to more frequent prior coryzal infections from seasonal coronaviruses. Alternatively, close contact with children may increase risk of SARs-CoV-2 infection. We investigated whether risk of infection with SARs-CoV-2 and severe outcomes differed between adults living with and without children.

**Methods:** Working on behalf of NHS England, we conducted a population-based cohort study using primary care data and pseudonymously-linked hospital and intensive care admissions, and death records, from patients registered in general practices representing 40% of England. Using multivariable Cox regression, we calculated fully-adjusted hazard ratios (HR) of outcomes from 1st February-3rd August 2020 comparing adults living with and without children in the household.

**Findings:** Among 9,157,814 adults ≤65 years, living with children 0-11 years was not associated with increased risks of recorded SARS-CoV-2 infection, COVID-19 related hospital or ICU admission but was associated with reduced risk of COVID-19 death (HR 0.75, 95%CI 0.62-0.92). Living with children aged 12-18 years was associated with a small increased risk of recorded SARS-CoV-2 infection (HR 1.08, 95%CI 1.03-1.13), but not associated with other COVID-19 outcomes. Living with children of any age was also associated with lower risk of dying from non-COVID-19 causes. Among 2,567,671 adults >65 years there was no association between living with children and outcomes related to SARS-CoV-2. We observed no consistent changes in risk following school closure.

**Interpretation:** For adults living with children there is no evidence of an increased risk of severe COVID-19 outcomes. These findings have implications for determining the benefit-harm balance of children attending school in the COVID-19 pandemic.

**Funding:** This work was supported by the Medical Research Council MR/V015737/1.

**Research in context:** *Evidence before this study:* We searched MEDLINE on 19th October 2020 for population-based epidemiological studies comparing the risk of SARS-CoV-2 infection and COVID-19 disease in people living with and without children. We searched for articles published in 2020, with abstracts available, and terms “(children or parents or dependants) AND (COVID or SARS-CoV-2 or coronavirus) AND (rate or hazard or odds or risk), in the title, abstract or keywords. 244 papers were identified for screening but none were relevant. One additional study in preprint was identified on medRxiv and found a reduced risk of hospitalisation for COVID-19 and a positive SARS-CoV-2 infection among adult healthcare workers living with children.

*Added value of this study:* This is the first population-based study to investigate whether the risk of recorded SARS-CoV-2 infection and severe outcomes from COVID-19 differ between adults living in households with and without school-aged children during the UK pandemic. Our findings show that for adults living with children there is no evidence of an increased risk of severe COVID-19 outcomes although there may be a slightly increased risk of recorded SARS-CoV-2 infection for working-age adults living with children aged 12 to 18 years. Working-age adults living with children 0 to 11 years have a lower risk of death from COVID-19 compared to adults living without children, with the effect size being comparable to their lower risk of death from any cause. We observed no consistent changes in risk of recorded SARS-CoV-2 infection and severe outcomes from COVID-19 comparing periods before and after school closure.

*Implications of all the available evidence:* Our results demonstrate no evidence of serious harms from COVID-19 to adults in close contact with children, compared to those living in households without children. This has implications for determining the benefit-harm balance of children attending school in the COVID-19 pandemic.

## Background

The role of children and adolescents in the transmission of SARS-CoV-2 is uncertain. They may have lower susceptibility to infection^1^ and are less likely to experience severe disease once infected.^2^ Modelling of other respiratory tract infections such as influenza suggests that children are a major driver of transmission during the initial phase of an epidemic, in part due to a high frequency of social contacts.^3^ However, accruing evidence suggests that for SARS-CoV-2, by contrast, lower susceptibility and possibly lower infectiousness among children means that they may not transmit infection more than adults.^4^

A suggested mechanism for lower susceptibility or propensity to disease is cross-protection due to immunity derived from recent seasonal coronavirus infection.^5^ There are four seasonal coronaviruses (hCoVs) that usually cause self-limiting “common cold”-like syndromes, although hCoVs are only one group of viruses responsible, causing 10 to 30% of common colds in adults.^6^ Children experience more colds each year than adults, with the highest infection frequency in young children.^7–9^ Adults in close contact with children also have a higher frequency of viral respiratory infections, especially adults exposed to younger children and among women.^10^

If recent hCoV infection is protective against SARS-CoV-2 infection or COVID-19, then adults living with children may be at a lower risk than those living without children. Conversely children may also introduce SARS-CoV-2 infections into their households, meaning adults living with children may experience an increased risk of infection and severe disease. In the face of increasing transmission in many countries and the need for policy decisions about school opening, quantifying the overall impact of living with children on the risk of SARS-CoV-2 infections and severe outcomes from COVID-19 is important. We therefore conducted a large cohort study using UK electronic health records (EHR) with linked data on household members to determine whether the risk of COVID-19 outcomes differs between adults living with and without school-aged children.

## Methods

### Database Description

Primary care records managed by the GP software provider The Phoenix Partnership (TPP) were linked to Secondary Uses Service (SUS) hospital admissions, Intensive Care National Audit & Research Centre (ICNARC) COVID-19-related Intensive Care Unit (ICU) admissions^11^ and Office for National Statistics (ONS) mortality records through OpenSAFELY, a data analytics platform created on behalf of NHS England to address urgent COVID-19 research questions (https://opensafely.org).

OpenSAFELY provides a secure software interface allowing the analysis of pseudonymized primary care patient records from England in near real-time within the EHR vendor’s highly secure data centre, avoiding the need for large volumes of potentially disclosive pseudonymized patient data to be transferred off-site. This, in addition to other technical and organisational controls, minimizes any risk of re-identification. Similarly, pseudonymized datasets from other data providers are securely provided to the EHR vendor and linked to the primary care data. The dataset analysed within OpenSAFELY is based on 24 million people currently registered with GP surgeries using TPP SystmOne software. It includes pseudonymized data such as coded diagnoses, medications and physiological parameters. No free text data are included. Further details on Information Governance and Ethics can be found on page 12.

### Study Design and Population

Our pre-specified study protocol^12^ and post-hoc protocol amendments (Table A1) are available.

The study population included all adults aged ≥18 years, registered and active for ≥three months, in an English TPP general practice on 1st February 2020 (study start). We followed participants until the earliest of: developing the outcome of interest; deregistration from their general practice; death from any cause; or 3rd August 2020 which was the latest date of linked outcomes, except for hospital admissions which were available until 1st May 2020.

### Inclusion & Exclusion Criteria

We required participants to have ≥three months of follow-up before study start to allow records to be updated following change of GP practice, while minimising loss of households moving home more frequently, potentially related to having children.

The TPP-developed pseudonymised household identifier links people living at the same address (Supplementary Methods). We excluded people with no household identifier, and individuals living in care homes (derived by TPP from linking addresses matched to publicly-available Care Quality Commission data) or household sizes ≥10 individuals (possible care homes or other institutions). Finally, we excluded households where any individual had a missing record of sex, age, or index of multiple deprivation (IMD), and we excluded individuals with missing ethnicity.

## Study Measures

### Exposures

The primary exposure was an ordered categorical variable reflecting school stages, derived using the ages of individuals linked by the household identifier; 1) no children under 18 years in the household, 2) any children 0 to 11 years of age in the household, 3) no children 0 to 11 years of age but one or more children aged 12 to 18 years in the household.

### Outcomes

We included four outcomes: 1) evidence of SARS-CoV-2 infection recorded in primary care defined as a code indicating either a clinical diagnosis of COVID-19, a positive swab test for SARS-CoV-2, or having sequelae of COVID-19 (Supplementary methods); 2) hospital admission for COVID-19 defined as a COVID-19 ICD-10 code in the primary diagnosis field (ascertained from SUS data); 3) ICU admission with COVID-19 that required non-invasive or invasive respiratory support (ascertained from ICNARC data); and 4) COVID-19 related death defined as a COVID-19 ICD-10 code anywhere on the death certificate (ascertained from ONS death certificate data). Participants were able to contribute to each outcome. *Post hoc*, to contextualise our findings, we added the outcome of non-COVID-19 death defined as death from any other cause on the death certificate.

### Covariates

We used a directed-acyclic graph approach to determining covariates (Figure A4), considering demographics including age in years, sex, body mass index (BMI; kg/m2), smoking status, IMD, ethnicity, geographic area, and the total number of adults in the household. We identified chronic comorbidities that are associated with the risk of severe COVID-19 outcomes.^13^ These are defined with links to code lists in the Supplementary Methods, with further information about demographic covariates. We identified participants who were recommended to shield following the UK government’s identification of clinically extremely vulnerable groups.^14^

## Statistical Analysis

### Primary Model

We analysed outcomes separately for adults aged 18-65 years, those most likely to be parents or primary caregivers, and also of working-age; and older adults (≥65 years). We described the proportion of individuals within each exposure and outcome, by the covariates. We then described the rate of outcomes according to the presence of children, and the number of children aged 0 to 11 years, in the household.

We used Cox proportional hazards modelling to determine hazard ratios (HRs) for each outcome using robust standard errors to account for clustering by household identifier and stratifying by geographic area to allow for regional variation in infection rates. We wished to adjust for illness in early adult life, which could have impacted the ability or decision to have children, as a confounder but were concerned about the accuracy of dates of onset of illness in the primary care record. Therefore, we used comorbidities at study start as a proxy for earlier health issues, as well as a marker of differences in current health status between adults living with and without children. To show the impact of this adjustment we present a ‘demographic-adjusted model’, adjusted for sex, age using a four-knot cubic spline, IMD, BMI, smoking, ethnicity and total number of adults in the household; and then a ‘comorbidity-adjusted model’ with addition of clinical comorbidities at study start. Violations of the proportional hazards assumption were explored by testing for a zero slope in the scaled Schoenfeld residuals.

### Secondary models

We examined possible interactions between our primary exposure and: 1) time periods from 1st February to 3rd April 2020 and 4th April 2020 to end of follow-up for each outcome. This period was chosen to allow three weeks after school closure on March 20th (except for key-worker children) for infections related to school transmission to progress to serious outcomes; 2) sex of the adult; and 3) probable shielding behaviour of the adult.

We also examined a potential ‘dose-response’ effect of exposure to previous coronavirus infections by recategorising the number of children between 0 and 11 years as 0 (reference), one, two, and ≥three.

### Sensitivity analyses

To confirm that our household identifier correctly linked parents/care-givers with children we conducted a similar analysis over an earlier time period (1st February 2019 to 1st February 2020) where the outcome was a Read code for threadworm infection, a condition where we anticipated transmission from young children to adults.

Secondly, we repeated the comorbidity-adjusted model: 1) restricting to participants with complete BMI and smoking data since in the main analysis we assumed those with missing BMI to be non-obese and those with missing smoking information to be non-smokers; 2) using age as the underlying timescale, to ensure we had fully adjusted for age as a confounder; 3) requiring ≥12 months primary care follow-up prior to study start, to fully capture pre-existing comorbidities; and 4) fitting time-interactions on covariates where there was evidence of non-proportional hazards.

Since data regarding occupation (and hence risk of SARS-CoV-2 infection) was not available, we conducted quantitative bias analysis to assess the potential extent of confounding from high-risk occupation among working-age adults. Bias-adjusted hazard ratios were calculated under a range of plausible assumptions regarding the association between occupation and risk of infection, and prevalence of high-risk occupations among those with and without children.^15–18^

Finally, we carried out an additional *post-hoc* analysis using multiple imputation to address those initially excluded for missing ethnicity data. Five imputed datasets were created with estimated hazard ratios combined using Rubin’s rules.^19^

## Software and Reproducibility

We used Python 3.8 and SQL (Server 2016 Enterprise SP2) for data management and Stata 16 for analysis. Analysis code is available online.^20^

## Results

The final cohort included 9,157,814 adults ≤65 years and 2,567,671 >65 years (Figure A1). Among those ≤65 years, 5,738,498 (63%) did not live with children, 2,568,901 (28%) lived with children 0 to 11 years and 9% (850,415) lived with children 12 to 18 years (Table 1). A total of 29,863 (0.33%) had evidence of SARS-CoV-2 infection recorded in their primary care record, 4,776 (0.05%) were admitted to hospital with COVID-19, 1,471 (0.02%) were admitted to ICU for ventilatory support with COVID-19 and 1,173 (0.01%) died of COVID-19 (Table A2). Those living with children were more likely to be younger, female, of non-white ethnicity, have a lower IMD, have more adults in the household and have fewer comorbidities.

**Table 1.** Cohort description, of adults 65 years and under, by presence of children or young people in the household

|  | **Total cohort, n (%)** | **No children under 18 years in the household (column %)** | **Children aged 0-11 years in the household (column %)** | **Children aged ≥12 years in the household (column %)** |
| --- | --- | --- | --- | --- |
| **Total** | 9157814 (100.0) | 5738498 (100.00) | 2568901 (100.00) | 850415 (100.00) |
| **Age** |  |  |  |  |
| 18-<30 | 1947216 (21.3) | 1339507 (23.34) | 447691 (17.43) | 160018 (18.82) |
| 30-<40 | 2202357 (24.0) | 1008794 (17.58) | 1105180 (43.02) | 88383 (10.39) |
| 40-<50 | 1991653 (21.7) | 877932 (15.30) | 778576 (30.31) | 335145 (39.41) |
| 50-<60 | 2012258 (22.0) | 1581524 (27.56) | 188351 (7.33) | 242383 (28.50) |
| 60-<66 | 1004330 (11.0) | 930741 (16.22) | 49103 (1.91) | 24486 (2.88) |
| **Sex** |  |  |  |  |
| Female | 4714908 (51.5) | 2778251 (48.41) | 1468018 (57.15) | 468639 (55.11) |
| Male | 4442906 (48.5) | 2960247 (51.59) | 1100883 (42.85) | 381776 (44.89) |
| **BMI (kg/m2)** |  |  |  |  |
| <18.5 | 178486 (1.9) | 119297 (2.08) | 46178 (1.80) | 13011 (1.53) |
| 18.5-24.9 | 2843580 (31.1) | 1791862 (31.23) | 819925 (31.92) | 231793 (27.26) |
| 25-29.9 | 2497554 (27.3) | 1545999 (26.94) | 717998 (27.95) | 233557 (27.46) |
| 30-34.9 (Obese class I) | 1252559 (13.7) | 787144 (13.72) | 343727 (13.38) | 121688 (14.31) |
| 35-39.9 (Obese class II) | 505136 (5.5) | 319669 (5.57) | 136592 (5.32) | 48875 (5.75) |
| ≥40 (Obese class III) | 270158 (3.0) | 173390 (3.02) | 70850 (2.76) | 25918 (3.05) |
| *Missing* | 1610341 (17.6) | 1001137 (17.45) | 433631 (16.88) | 175573 (20.65) |
| **Smoking** |  |  |  |  |
| Never | 4405852 (48.1) | 2757196 (48.05) | 1234664 (48.06) | 413992 (48.68) |
| Former | 2636324 (28.8) | 1629388 (28.39) | 760180 (29.59) | 246756 (29.02) |
| Current | 1869068 (20.4) | 1208623 (21.06) | 510614 (19.88) | 149831 (17.62) |
| *Missing* | 246570 (2.7) | 143291 (2.50) | 63443 (2.47) | 39836 (4.68) |
| **Ethnicity** |  |  |  |  |
| White | 7569454 (82.7) | 4901365 (85.41) | 1986318 (77.32) | 681771 (80.17) |
| Mixed | 158349 (1.7) | 94318 (1.64) | 49785 (1.94) | 14246 (1.68) |
| South Asian | 832720 (9.1) | 393138 (6.85) | 340500 (13.25) | 99082 (11.65) |
| Black | 306539 (3.3) | 166789 (2.91) | 106666 (4.15) | 33084 (3.89) |
| Other | 290752 (3.2) | 182888 (3.19) | 85632 (3.33) | 22232 (2.61) |
| **IMD quintile** |  |  |  |  |
| 1 (least deprived) | 1684917 (18.4) | 1033896 (18.02) | 470250 (18.31) | 180771 (21.26) |
| 2 | 1805617 (19.7) | 1165046 (20.30) | 473081 (18.42) | 167490 (19.70) |
| 3 | 1874964 (20.5) | 1219743 (21.26) | 491526 (19.13) | 163695 (19.25) |
| 4 | 1976632 (21.6) | 1250521 (21.79) | 554876 (21.60) | 171235 (20.14) |
| 5 (most deprived) | 1815684 (19.8) | 1069292 (18.63) | 579168 (22.55) | 167224 (19.66) |
| **Total number adults in household** |  |  |  |  |
| 1 | 2226830 (24.3) | 1750754 (30.51) | 376825 (14.67) | 99251 (11.67) |
| 2 | 3779868 (41.3) | 2013500 (35.09) | 1421964 (55.35) | 344404 (40.50) |
| □3 | 3151116 (34.4) | 1974244 (34.40) | 770112 (29.98) | 406760 (47.83) |
| **Blood pressure** |  |  |  |  |
| Normal | 2440215 (26.6) | 1347688 (23.49) | 866256 (33.72) | 226271 (26.61) |
| Elevated | 1324098 (14.5) | 814512 (14.19) | 390162 (15.19) | 119424 (14.04) |
| High Stage 1 | 2878905 (31.4) | 1844169 (32.14) | 759199 (29.55) | 275537 (32.40) |
| High Stage 2 | 1549272 (16.9) | 1082545 (18.86) | 320825 (12.49) | 145902 (17.16) |
| Missing | 965324 (10.5) | 649584 (11.32) | 232459 (9.05) | 83281 (9.79) |
| **High bp or diagnosed hypertension** | 2156211 (23.5) | 1548682 (26.99) | 409471 (15.94) | 198058 (23.29) |
| **Comorbidities** |  |  |  |  |
| **Chronic respiratory disease ex asthma** | 184872 (2.0) | 150741 (2.63) | 21629 (0.84) | 12502 (1.47) |
| **Asthma** | 1574912 (17.2) | 977849 (17.04) | 448542 (17.46) | 148521 (17.46) |
| **Chronic cardiac disease** | 259057 (2.8) | 203191 (3.54) | 35929 (1.40) | 19937 (2.34) |
| **Diabetes** |  |  |  |  |
| No diabetes | 8712703 (95.1) | 5412597 (94.32) | 2489806 (96.92) | 810300 (95.28) |
| Type 1, controlled | 12483 (0.1) | 8459 (0.15) | 3089 (0.12) | 935 (0.11) |
| Type 1, uncontrolled | 37382 (0.4) | 25807 (0.45) | 8412 (0.33) | 3163 (0.37) |
| Type 2, controlled | 219161 (2.4) | 162903 (2.84) | 36861 (1.43) | 19397 (2.28) |
| Type 2, uncontrolled | 171932 (1.9) | 126000 (2.20) | 29659 (1.15) | 16273 (1.91) |
| Diabetes, no HbA1c | 4153 (0.0) | 2732 (0.05) | 1074 (0.04) | 347 (0.04) |
| **Haematological cancer** |  |  |  |  |
| Diagnosed < 1 year ago | 2349 (0.0) | 1733 (0.03) | 420 (0.02) | 196 (0.02) |
| Diagnosed 1-4.9 years ago | 7332 (0.1) | 5538 (0.10) | 1211 (0.05) | 583 (0.07) |
| Diagnosed ≥5 years ago | 19225 (0.2) | 14017 (0.24) | 3595 (0.14) | 1613 (0.19) |
| **Non-haematological cancer** |  |  |  |  |
| Diagnosed < 1 year ago | 22171 (0.2) | 16737 (0.29) | 3454 (0.13) | 1980 (0.23) |
| Diagnosed 1-4.9 years ago | 64176 (0.7) | 48734 (0.85) | 9648 (0.38) | 5794 (0.68) |
| Diagnosed ≥5 years ago | 113243 (1.2) | 89421 (1.56) | 14406 (0.56) | 9416 (1.11) |
| **Reduced kidney function** |  |  |  |  |
| Estimated GFR 30-60 | 93017 (1.0) | 77514 (1.35) | 9300 (0.36) | 6203 (0.73) |
| Estimated GFR <30 | 9067 (0.1) | 7063 (0.12) | 1298 (0.05) | 706 (0.08) |
| **End-stage renal disease\*** | 10815 (0.1) | 8153 (0.14) | 1827 (0.07) | 835 (0.10) |
| **Chronic Liver disease** | 51404 (0.6) | 39107 (0.68) | 8860 (0.34) | 3437 (0.40) |
| **Stroke/dementia** | 71425 (0.8) | 57346 (1.00) | 9030 (0.35) | 5049 (0.59) |
| **Other neurological disease** | 63572 (0.7) | 48570 (0.85) | 9977 (0.39) | 5025 (0.59) |
| **Solid organ transplant\*\*** | 2103 (0.0) | 1612 (0.03) | 317 (0.01) | 174 (0.02) |
| **Asplenia** | 10867 (0.1) | 7818 (0.14) | 2080 (0.08) | 969 (0.11) |
| **Rheumatoid/Lupus/ Psoriasis** | 409631 (4.5) | 265742 (4.63) | 104306 (4.06) | 39583 (4.65) |
| **Other immunosuppressive condition** | 32745 (0.4) | 22907 (0.40) | 7280 (0.28) | 2558 (0.30) |
| **Probable shielding\*\*\*** | 1922340 (21.0) | 1248677 (21.76) | 497917 (19.38) | 175746 (20.67) |
| **Any comorbidity\*\*\*\*** | 2747911 (30.0) | 1824629 (31.80) | 671397 (26.14) | 251885 (29.62) |

\*End-stage renal disease includes on dialysis or having had a kidney transplant
\*\*All solid organ transplants, excluding kidney
\*\*\*Shielding includes organ transplant recipients, renal replacement therapy, haematological cancers, non-haematological cancers, immunodeficiencies/asplenia and severe respiratory conditions.
\*\*\*\*Any comorbidity includes: chronic respiratory disease, asthma, chronic cardiac disease, diabetes, cancer, end-stage renal disease, chronic liver disease, stroke or dementia, other neurological disease, other transplant, asplenia, Rheumatoid/Lupus/ Psoriasis or other immunosuppressive condition

Among those >65 years, 2,481,210 (97%) did not live with children (Table 2). A total of 11,826 (0.46%) had evidence of SARS-CoV-2 infection recorded in their primary care record, 6,496 (0.25%) were admitted to hospital with COVID-19, 675 (0.03%) were admitted to ICU for ventilatory support with COVID-19 and 6,352 (0.25%) died of COVID-19 (Table A3).

**Table 2.** Cohort description, of adults over 65 years, by presence of children or young people in the household

|  | Total cohort, n (%) | No children under 18 years in the household (column %) | Children aged 0-11 years in the household (column %) | Children aged ≥12 years in the household (column %) |
| --- | --- | --- | --- | --- |
| **Total** | 2567671 (100.0) | 2481210 (100.00) | 58165 (100.00) | 28296 (100.00) |
| **Age** |  |  |  |  |
| >65-70 | 609634 (23.7) | 578189 (23.30) | 22682 (39.00) | 8763 (30.97) |
| 70-80 | 1300326 (50.6) | 1258494 (50.72) | 27598 (47.45) | 14234 (50.30) |
| 80+ | 657711 (25.6) | 644527 (25.98) | 7885 (13.56) | 5299 (18.73) |
| **Sex** |  |  |  |  |
| Female | 1384514 (53.9) | 1338333 (53.94) | 31332 (53.87) | 14849 (52.48) |
| Male | 1183157 (46.1) | 1142877 (46.06) | 26833 (46.13) | 13447 (47.52) |
| **BMI (kg/m2)** |  |  |  |  |
| <18.5 | 44163 (1.7) | 42958 (1.73) | 789 (1.36) | 416 (1.47) |
| 18.5-24.9 | 760180 (29.6) | 738360 (29.76) | 14883 (25.59) | 6937 (24.52) |
| 25-29.9 | 949715 (37.0) | 919015 (37.04) | 20418 (35.10) | 10282 (36.34) |
| 30-34.9 (Obese class I) | 462119 (18.0) | 445225 (17.94) | 11400 (19.60) | 5494 (19.42) |
| 35-39.9 (Obese class II) | 151009 (5.9) | 145127 (5.85) | 3854 (6.63) | 2028 (7.17) |
| ≥40 (Obese class III) | 58570 (2.3) | 56065 (2.26) | 1681 (2.89) | 824 (2.91) |
| *Missing* | 141915 (5.5) | 134460 (5.42) | 5140 (8.84) | 2315 (8.18) |
| **Smoking** |  |  |  |  |
| Never | 1011539 (39.4) | 969700 (39.08) | 29086 (50.01) | 12753 (45.07) |
| Former | 1336686 (52.1) | 1301772 (52.47) | 22475 (38.64) | 12439 (43.96) |
| Current | 213854 (8.3) | 205070 (8.26) | 5893 (10.13) | 2891 (10.22) |
| *Missing* | 5592 (0.2) | 4668 (0.19) | 711 (1.22) | 213 (0.75) |
| **Ethnicity** |  |  |  |  |
| White | 2419165 (94.2) | 2362620 (95.22) | 36022 (61.93) | 20523 (72.53) |
| Mixed | 9797 (0.4) | 8670 (0.35) | 760 (1.31) | 367 (1.30) |
| South Asian | 90017 (3.5) | 67805 (2.73) | 16729 (28.76) | 5483 (19.38) |
| Black | 27290 (1.1) | 23616 (0.95) | 2531 (4.35) | 1143 (4.04) |
| Other | 21402 (0.8) | 18499 (0.75) | 2123 (3.65) | 780 (2.76) |
| **IMD quintile** |  |  |  |  |
| 1 (least deprived) | 618192 (24.1) | 602937 (24.30) | 9811 (16.87) | 5444 (19.24) |
| 2 | 596748 (23.2) | 580549 (23.40) | 10773 (18.52) | 5426 (19.18) |
| 3 | 548147 (21.3) | 530203 (21.37) | 11944 (20.53) | 6000 (21.20) |
| 4 | 460588 (17.9) | 441284 (17.79) | 13271 (22.82) | 6033 (21.32) |
| 5 (most deprived) | 343996 (13.4) | 326237 (13.15) | 12366 (21.26) | 5393 (19.06) |
| **Total number adults in household** |  |  |  |  |
| 1 | 877922 (34.2) | 874751 (35.26) | 1663 (2.86) | 1508 (5.33) |
| 2 | 1319765 (51.4) | 1303437 (52.53) | 9341 (16.06) | 6987 (24.69) |
| ≥3 | 369984 (14.4) | 303022 (12.21) | 47161 (81.08) | 19801 (69.98) |
| **Blood pressure** |  |  |  |  |
| Normal | 298019 (11.6) | 287358 (11.58) | 7296 (12.54) | 3365 (11.89) |
| Elevated | 400138 (15.6) | 386920 (15.59) | 8935 (15.36) | 4283 (15.14) |
| High Stage 1 | 947144 (36.9) | 915718 (36.91) | 21174 (36.40) | 10252 (36.23) |
| High Stage 2 | 909480 (35.4) | 880644 (35.49) | 18968 (32.61) | 9868 (34.87) |
| Missing | 12890 (0.5) | 10570 (0.43) | 1792 (3.08) | 528 (1.87) |
| **High bp or diagnosed hypertension** | 1769252 (68.9) | 1711606 (68.98) | 38244 (65.75) | 19402 (68.57) |
| **Comorbidities** |  |  |  |  |
| **Chronic respiratory disease ex asthma** | 317732 (12.4) | 308239 (12.42) | 6048 (10.40) | 3445 (12.17) |
| **Asthma** | 347416 (13.5) | 334967 (13.50) | 8395 (14.43) | 4054 (14.33) |
| **Chronic cardiac disease** | 536239 (20.9) | 518877 (20.91) | 11355 (19.52) | 6007 (21.23) |
| **Diabetes** |  |  |  |  |
| No diabetes | 2103299 (81.9) | 2040001 (82.22) | 42106 (72.39) | 21192 (74.89) |
| Type 1, controlled | 3270 (0.1) | 3174 (0.13) | 62 (0.11) | 34 (0.12) |
| Type 1, uncontrolled | 6772 (0.3) | 6580 (0.27) | 119 (0.20) | 73 (0.26) |
| Type 2, controlled | 311147 (12.1) | 296882 (11.97) | 9798 (16.85) | 4467 (15.79) |
| Type 2, uncontrolled | 141345 (5.5) | 132921 (5.36) | 5944 (10.22) | 2480 (8.76) |
| Diabetes, no HbA1c | 1838 (0.1) | 1652 (0.07) | 136 (0.23) | 50 (0.18) |
| **Haematological cancer** |  |  |  |  |
| Diagnosed < 1 year ago | 3661 (0.1) | 3563 (0.14) | 62 (0.11) | 36 (0.13) |
| Diagnosed 1-4.9 years ago | 11073 (0.4) | 10776 (0.43) | 189 (0.32) | 108 (0.38) |
| Diagnosed ≥5 years ago | 22672 (0.9) | 22089 (0.89) | 387 (0.67) | 196 (0.69) |
| **Non-haematological cancer** |  |  |  |  |
| Diagnosed < 1 year ago | 30875 (1.2) | 30042 (1.21) | 557 (0.96) | 276 (0.98) |
| Diagnosed 1-4.9 years ago | 91267 (3.6) | 88872 (3.58) | 1578 (2.71) | 817 (2.89) |
| Diagnosed ≥5 years ago | 243542 (9.5) | 238178 (9.60) | 3378 (5.81) | 1986 (7.02) |
| **Reduced kidney function** |  |  |  |  |
| Estimated GFR 30-60 | 535850 (20.9) | 521234 (21.01) | 9334 (16.05) | 5282 (18.67) |
| Estimated GFR <30 | 39509 (1.5) | 38103 (1.54) | 939 (1.61) | 467 (1.65) |
| **End-stage renal disease\*** | 6279 (0.2) | 5922 (0.24) | 249 (0.43) | 108 (0.38) |
| **Chronic Liver disease** | 24604 (1.0) | 23696 (0.96) | 609 (1.05) | 299 (1.06) |
| **Stroke/dementia** | 187946 (7.3) | 182242 (7.34) | 3733 (6.42) | 1971 (6.97) |
| **Other neurological disease** | 50577 (2.0) | 49058 (1.98) | 1018 (1.75) | 501 (1.77) |
| **Solid organ transplant\*\*** | 825 (0.0) | 774 (0.03) | 34 (0.06) | 17 (0.06) |
| **Asplenia** | 6355 (0.2) | 6190 (0.25) | 99 (0.17) | 66 (0.23) |
| **Rheumatoid/Lupus/ Psoriasis** | 199014 (7.8) | 193236 (7.79) | 3755 (6.46) | 2023 (7.15) |
| **Other immunosuppressive condition** | 5235 (0.2) | 5030 (0.20) | 135 (0.23) | 70 (0.25) |
| **Probable shielding\*\*\*** | 903291 (35.2) | 876412 (35.32) | 17581 (30.23) | 9298 (32.86) |
| **Any comorbidity\*\*\*\*** | 1554606 (60.5) | 1502301 (60.55) | 34800 (59.83) | 17505 (61.86) |

\*End-stage renal disease includes on dialysis or having had a kidney transplant
\*\*All solid organ transplants, excluding kidney
\*\*\*Shielding includes organ transplant recipients, renal replacement therapy, haematological cancers, non-haematological cancers, immunodeficiencies/asplenia and severe respiratory conditions.
\*End-stage renal disease includes on dialysis or having had a kidney transplant
\*\*All solid organ transplants, excluding kidney
\*\*\*\*Any comorbidity includes: chronic respiratory disease, asthma, chronic cardiac disease, diabetes, cancer, end-stage renal disease, chronic liver disease, stroke or dementia, other neurological disease, other transplant, asplenia, Rheumatoid/Lupus/ Psoriasis or other immunosuppressive condition

Among adults ≤65 years, after adjusting for ethnicity, IMD, BMI, smoking and total number of adults in the household, living with children aged 0 to 11 years was not associated with recorded SARS-CoV-2 infection, COVID-19 related hospital or ICU admission but was associated with a reduced risk of death from COVID-19 (HR 0.74, 95%CI 0.60-0.90) (Table A4). Living with children aged 12 to 18 years was associated with a small increased risk of recorded SARS-CoV-2 infection (HR 1.08, 95%CI 1.03-1.13), but was not associated with COVID-19 related hospital or ICU admission, or death from COVID-19 (Table A4).

For adults >65 years living in a household with children, there was no evidence of an association with any outcome (Table A4).

Living with children of any age was associated with around 30% reduced risk of death from non-COVID-19 causes for adults ≤65 but there was no reduction in risk for adults >65 years (Table A4).

In all analyses, additionally adjusting for comorbidities did not materially change the results (Figure 1, Table A4).

**Figure 1:**
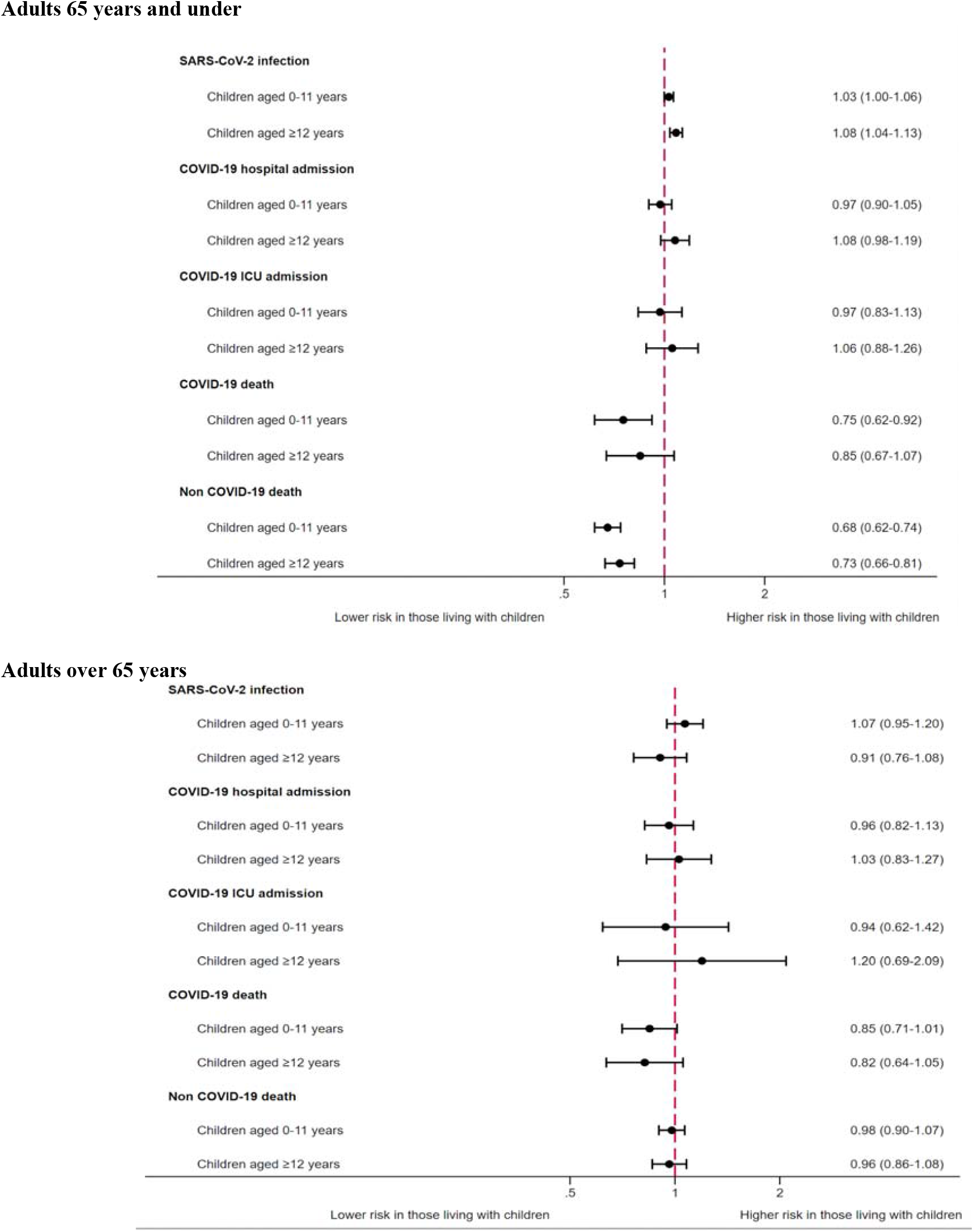
Adjusted* Hazard Ratios (HRs) for outcomes ((a) recorded SARS-CoV-2 infection, (b) COVID-19 hospital admission, (c) COVID-19 ICU admission, (d) COVID-19 death and (e) non-COVID-19 death), stratified by age. *Comorbidity-adjusted model: Adjusted for age, sex, ethnicity, number adults in household, IMD, BMI, smoking, hypertension or high blood pressure, chronic respiratory disease, asthma, cancer, chronic liver disease, stroke or dementia, other neurological disease, reduced kidney function, end-stage renal disease, solid organ transplant, asplenia, rheumatoid, lupus or psoriasis, other immunosuppressive condition.

There were no evident trends in the associations between risks of recorded SARS-CoV-2 in primary care or severe outcomes from COVID-19 and the number of children aged 0 to 11 years in a household, for adults of any age (Table A4).

We explored whether the association between household exposure to children and the risk of COVID-19 disease varied by sex, time period in relation to school closure, and probable shielding status, in the comorbidity-adjusted model (Figure 2 for adults ≤65 years and Supplementary Figure A5 for adults ≥65 years).

**Figure 2.**
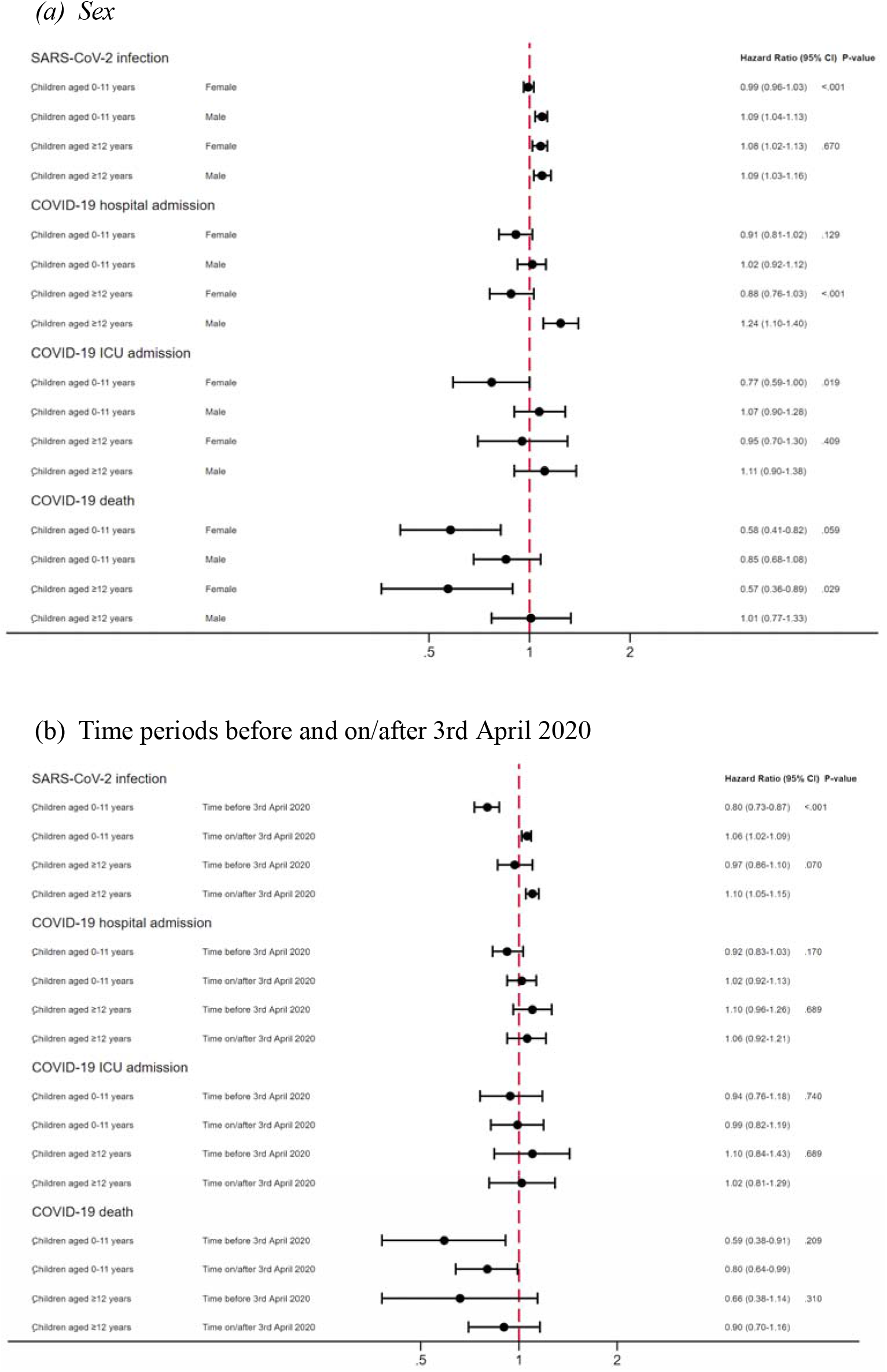
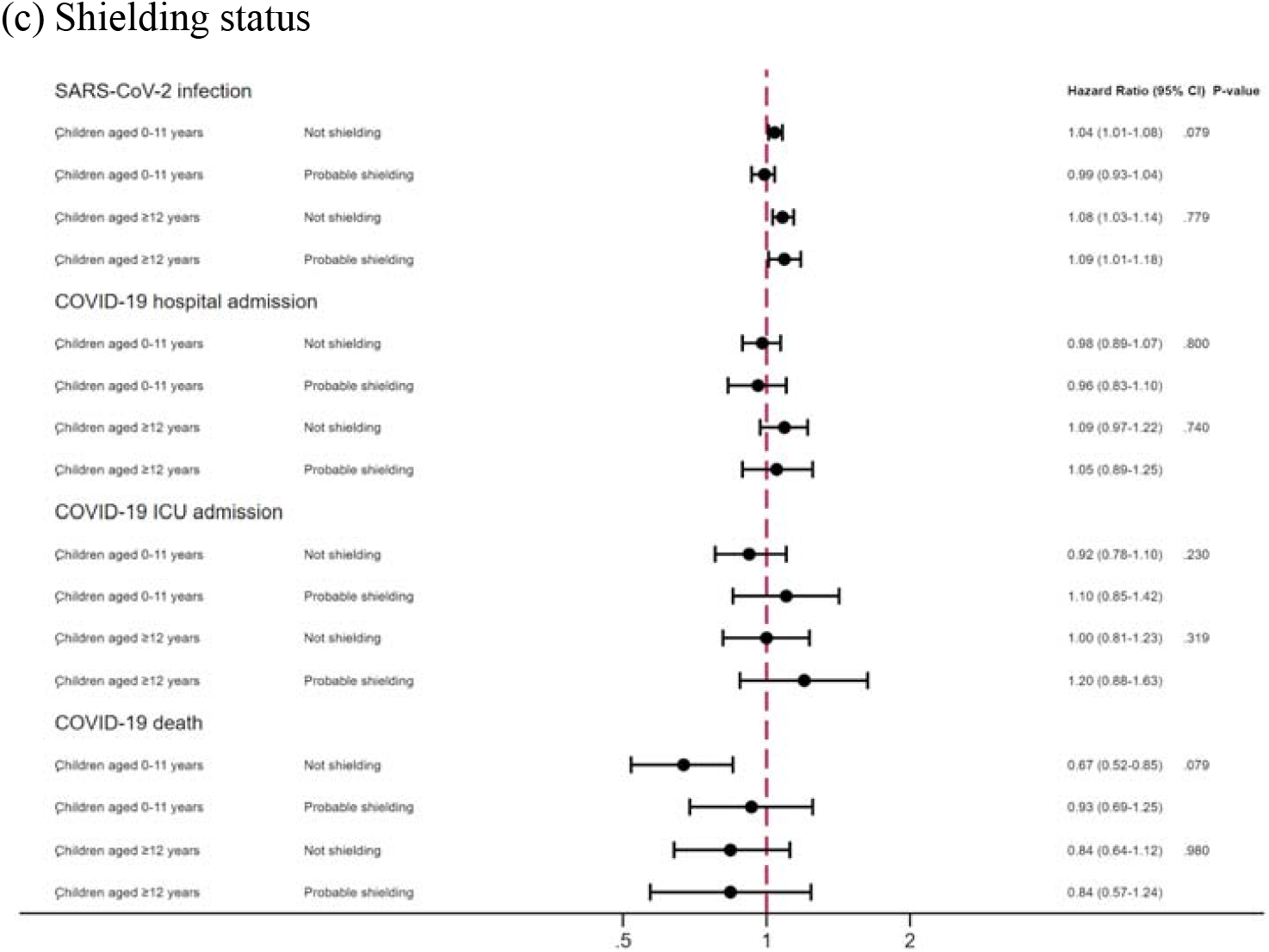
Comorbidity adjusted hazard ratios (HRs) and 95% confidence intervals (CI) for each COVID-19 outcome, compared to having no children in the household by (a) sex, (b) time periods before and after 3rd April 2020 and (c) shielding status among those 65 years and under.

Among ≤65 year olds, there was some evidence that the associations varied by sex of the adult: for recorded SARS-CoV-2 infection (P-value for interaction=<0.001), the small increased risk among those living with children aged 0 to 11 years was observed in males (HR 1.09, 95% CI 1.04-1.13) but not females (HR 0.99, 95%CI 0.96-1.03). For COVID-19-related hospital admission (P int=<0.001), there was evidence of an increased risk of admission among males living with children aged 12 to 18 years (HR 1.24, 95%CI 1.10-1.40), but not females (HR 0.88, 95% CI 0.76-1.03). For death from COVID-19 there was evidence of a reduced risk in females (HR 0.58, 95%CI 0.41-0.82 for females living with children 0 to 11 years (P int=0.059); HR 0.57, 95% CI 0.36-0.89 (P int=0.029) for those living with children 12 to 18 years), but no evidence of lower risk for males living with children.

There was evidence that for working-age adults, the risk of recorded SARS-CoV-2 infection was higher in the period three weeks after school closures compared to earlier but no evidence of important variations in the risk of any other COVID-19 outcomes between these time periods. There was no evidence of important increases in risk for any outcome for working-age adults who were likely to have been shielding and no evidence of any interactions for those aged ≥65 years.

### Model checking and sensitivity analyses

Among adults ≤65 years, living with children aged 0 to 11 years was associated with a 2-fold increased risk of being diagnosed with threadworm (HR 2.53, 95%CI 2.19-2.92), with strong evidence of an increased risk with increasing number of children in the household, and an increased risk among adults living with children aged 12 to 18 years, though of a smaller magnitude (HR 1.32, 95%CI 1.03-1.70) (Table A4).

None of the sensitivity analyses materially altered the results from the comorbidity adjusted models (Figure A3).

When accounting for missing ethnicity data through multiple imputation, the associations between living with children and outcomes remained the same as the primary (complete case) analysis (Table A6). Quantitative bias analysis accounting for shared risk of people with young children working in occupations with high-risk of exposure did not materially alter any results (Tables A7-A14).

## Discussion

We observed no increased risk of recorded infection or serious COVID-19 outcomes among working-age adults sharing a household with children aged 0 to 11 years, compared to those living without children, but a reduced risk of death from COVID-19. This was similar in magnitude to the reduction in risk of death from causes other than COVID-19 seen in working-age adults living with children of all ages. For working-age adults living with children aged 12 to 18 years we found a small (between three and 13%) increased risk of recorded SARS-CoV-2 infection, but no association with other outcomes from COVID-19. We found no association with the risk of recorded SARS-CoV-2 infection and outcomes from COVID-19 among adults over 65 years living with children of any age.

Our analysis has a number of important strengths. Firstly, our study is large, providing us with sufficient power to examine rarer outcomes, as well as interactions with several important factors. Secondly we show largely consistent results across SARS-CoV-2 infection and COVID-19 related clinical outcomes, supporting our findings. Thirdly, in our control analysis of threadworm infection in adults we show strong evidence that our model can detect an infection transmitted from children to adults, with, as we would anticipate, risk increasing with the number of children in a household and a stronger effect seen in younger children.

However, there are also limitations. During the period covered by this study, the outcome of recorded SARS-CoV-2 infection is mainly based on swab tests taken in community testing centres and healthcare settings that were later transferred to the primary care record. Therefore, results related to testing should be viewed as likely to be heavily influenced by people in high-risk jobs where testing was more easily available. A positive recorded infection combines the risk of being infected with the chance of being tested and this is particularly important for interpreting the interaction by date. Occupation was also an unmeasured confounder both in terms of exposure to SARS-CoV-2 (such as healthcare and other high-risk workers) and degree of contact with children outside the home (such as nursery workers). To explore the potential impact of lack of occupational information we conducted a quantitative bias analysis which did not meaningfully change our results for any outcome. We were not able to adjust for confounding by previous comorbidities that affected ability or choice to have children, and subsequent risk of development of severe outcomes from COVID-19. However, to examine the potential impact of this we show results from models with and without adjusting for baseline comorbidities and again find no important differences. It is likely we have misclassified the degree of contact with children in a number of situations such as for divorced parents, and limitations in the data may mean misclassification of the number of people living in a household, e.g. for flats within a larger property or when people have not updated their address with their general practice following a house move. Finally, in this analysis we are assuming a constant relationship for infections between people through clustering at the household level, rather than detailed modelling of how infections are transmitted within households.

One previous cohort study of 310,097 healthcare workers and other adults in their household in Scotland has also addressed whether the risk of SARS-CoV-2 infection and severe outcomes from COVID-19 differs between adults living in households with and without children aged 0 to 11 years.^21^ Similar to our results they find no increase in hospitalisation due to COVID-19 for people living with young children. However, they find a slightly reduced rate of testing positive for COVID-19 for adults living with young children, and evidence that the protective effect increased with a greater number of children. Differences in power, greater consistency in testing patterns and exposure risk among healthcare workers in their cohort and different epidemic trajectories between Scotland and England make direct comparisons between our results difficult.

Our findings of no increase in risk of severe outcomes from COVID-19 despite some evidence of a small increased risk of infection among adults living with older children could be explained by confounding if parents are healthier than people without children. This explanation is supported by our finding of a substantially lower risk of mortality from causes other than COVID-19 among working-age adults living with children of any age. Parents are known to have lower all-cause mortality than individuals without children.^22,23^ The protective mechanisms of having children are likely to be multifactorial, including healthier behaviours among parents, e.g. in relation to smoking and alcohol,^24,25^ and self-selection of healthier individuals becoming parents.^26^ However, beneficial changes in immune function from exposure to young children have been proposed to cause reduced mortality among parents.^27^

In terms of implications for health policy, our results can be viewed as the cumulative effects of transmission risk of SARS-CoV-2, the impact of any cross-reactive immunity and differences in underlying health status and health related behaviours of adults living with and without children. We found that outcomes differed by the sex of the adult, with in general, less risk of severe outcomes and lower risk of death for women compared to men, particularly those living with younger children. Given that women and parents of younger children have higher rates of respiratory tract infections,^10^ this offers some clinical support to the basic science evidence that exposure to other hCoVs may offer some immunity to SARS-CoV-2.^28^ If confirmed, this would have important implications for understanding high-risk populations and the likely future course of the pandemic.

Concern that children act as an important source of spread of SARS-CoV-2 have led to school closures in many countries during the pandemic. We saw an increased risk of recorded SARS-CoV-2 infection for adults living with children in the period from three weeks after schools closed, compared to previously, although this must be interpreted in light of limitations of testing data. However, overall our findings suggest that on a population level transmission from school age children does not result in an increased risk of serious outcomes among the adults they live with.

## Conclusion

During a period covering the first peak of the UK pandemic of SARS-CoV-2 we found no evidence of increased risk of serious outcomes from COVID-19 among adults of any age living with children compared to those in households without children. Within the limitations of testing data there was evidence of a small increased risk of recorded SARS-CoV-2 infection for adults living with older children. We found no evidence for a reduction in risk following school closure. These findings, in consideration alongside other evidence, have implications for determining the benefit-harm balance of children attending school in the COVID-19 pandemic.

## Data Availability

All data were linked, stored, and analysed securely within the OpenSAFELY platform. Detailed pseudonymised patient data are potentially re-identifiable and therefore not shared. We rapidly delivered the OpenSAFELY data analysis platform without previous funding to deliver timely analyses of urgent research questions in the context of the global COVID-19 health emergency: now that the platform is established, we are developing a formal process for external users to request access in collaboration with NHS England.

## Acknowledgements

We are very grateful for all the support received from the TPP Technical Operations team throughout this work, and for generous assistance from the information governance and database teams at NHS England / NHSX.

## Information governance and ethical approval

NHS England is the data controller; TPP is the data processor; and the key researchers on OpenSAFELY are acting on behalf of NHS England. This implementation of OpenSAFELY is hosted within the TPP environment which is accredited to the ISO 27001 information security standard and is NHS IG Toolkit compliant;^29,30^ patient data has been pseudonymised for analysis and linkage using industry standard cryptographic hashing techniques; all pseudonymised datasets transmitted for linkage onto OpenSAFELY are encrypted; access to the platform is via a virtual private network (VPN) connection, restricted to a small group of researchers; the researchers hold contracts with NHS England and only access the platform to initiate database queries and statistical models; all database activity is logged; only aggregate statistical outputs leave the platform environment following best practice for anonymisation of results such as statistical disclosure control for low cell counts.^31^ The OpenSAFELY research platform adheres to the data protection principles of the UK Data Protection Act 2018 and the EU General Data Protection Regulation (GDPR) 2016. In March 2020, the Secretary of State for Health and Social Care used powers under the UK Health Service (Control of Patient Information) Regulations 2002 (COPI) to require organisations to process confidential patient information for the purposes of protecting public health, providing healthcare services to the public and monitoring and managing the COVID-19 outbreak and incidents of exposure.^32^ Taken together, these provide the legal bases to link patient datasets on the OpenSAFELY platform. GP practices, from which the primary care data are obtained, are required to share relevant health information to support the public health response to the pandemic, and have been informed of the OpenSAFELY analytics platform.

This study was approved by the Health Research Authority (REC reference 20/LO/0651) and by the LSHTM Ethics Board (reference 21863).

### Guarantor

HF/LT/LS/BG are guarantors

### Contributorship

Conceptualization: HF, LT, CEM, HIM, CM, JPB, LS, SJWE, KB, RME, BG

Data curation: HF, KB, CEM, EJW, AS, RM, CTR, AJW, CB, JC, CM, WJH, BM, SB, HJC, RM

Formal analysis: HF, KB, JPB

Funding acquisition: LS, BG, RME

Investigation: HF, LT, CEM, SB

Methodology: HF, LT, RM, AS, CTR, KB, KW, RME, LS, BG, BM, EJW, HJC, HIM, SJWE, CEM, JPB

Codelists: HF, RM, LT, AS, AJW, CM, BG, WJH, SB, AM, CEM, HD, KW

Project administration: HF, LT, AS, AJW, CEM, BG, WJH

Resources: CB JC BG BM SB AM

Software: CB JC DE PI CEM WJH SB HJC RC JP FH SH ND

Visualisation: HF, SB

Writing - original draft: HF, LT

Writing-review & editing: ALL

Information governance: CB LS BG AM

Verification of the underlying data: HF, CEM, KB, CB and JC

