## Supplementary material for "Association between living with children and outcomes from COVID-19: an OpenSAFELY cohort study of 12 million adults in England": Data supplement

### **Appendix**

**Figure A1: Flow diagram of cohort with numbers excluded at different stages**

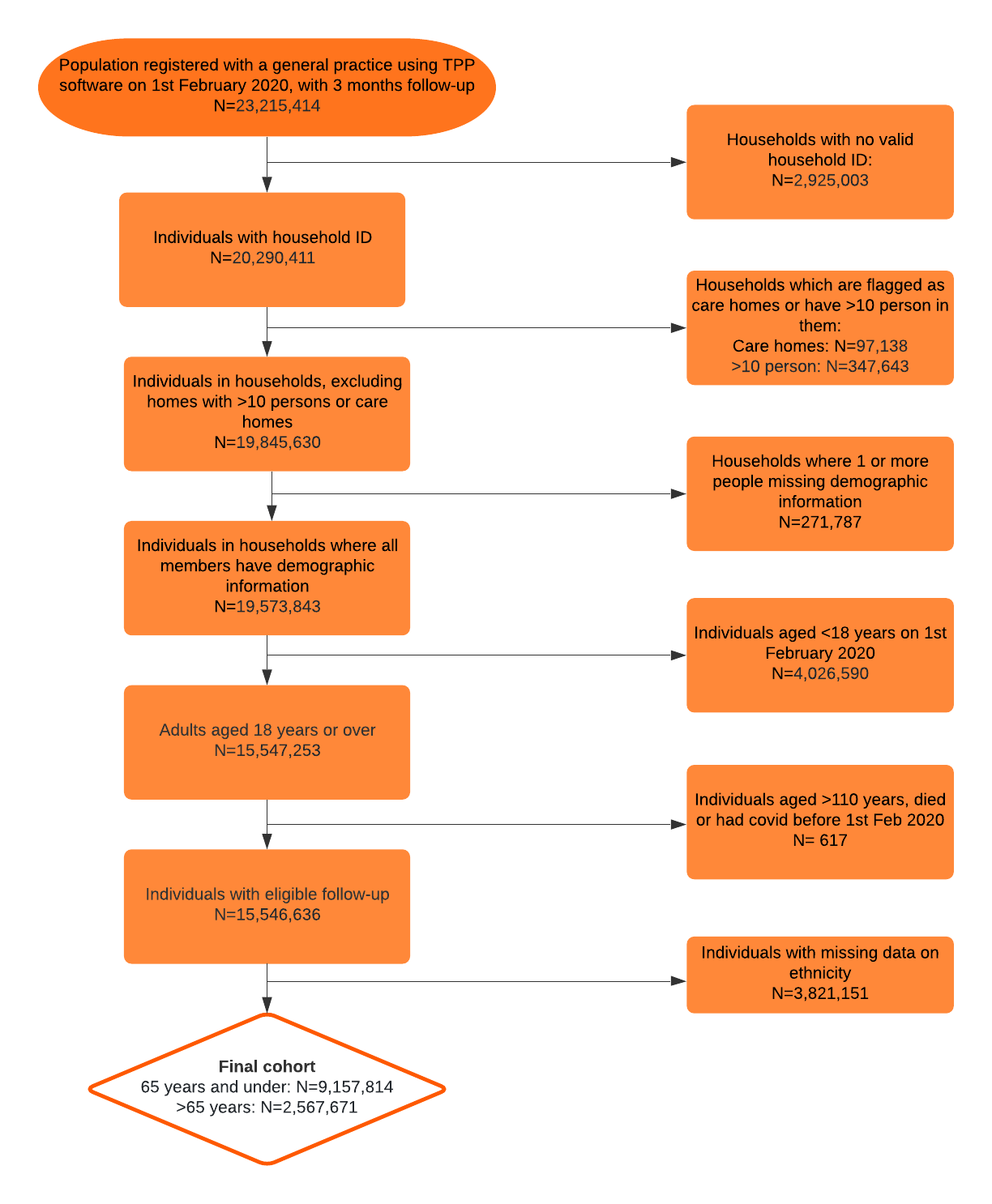
<https://app.lucidchart.com/documents/edit/7611be3a-feba-488e-8ace-b4f1b9ee44e2/0?callback=close&name=docs&callback_type=back&v=1740&s=612>

### **Figure A2: Histograms of frequencies of outcomes (evidence of SARS-CoV-2 infection recorded in primary care, COVID-19 outcomes and non-COVID-19 deaths) over the study period, 1st February to 3rd August 2020**

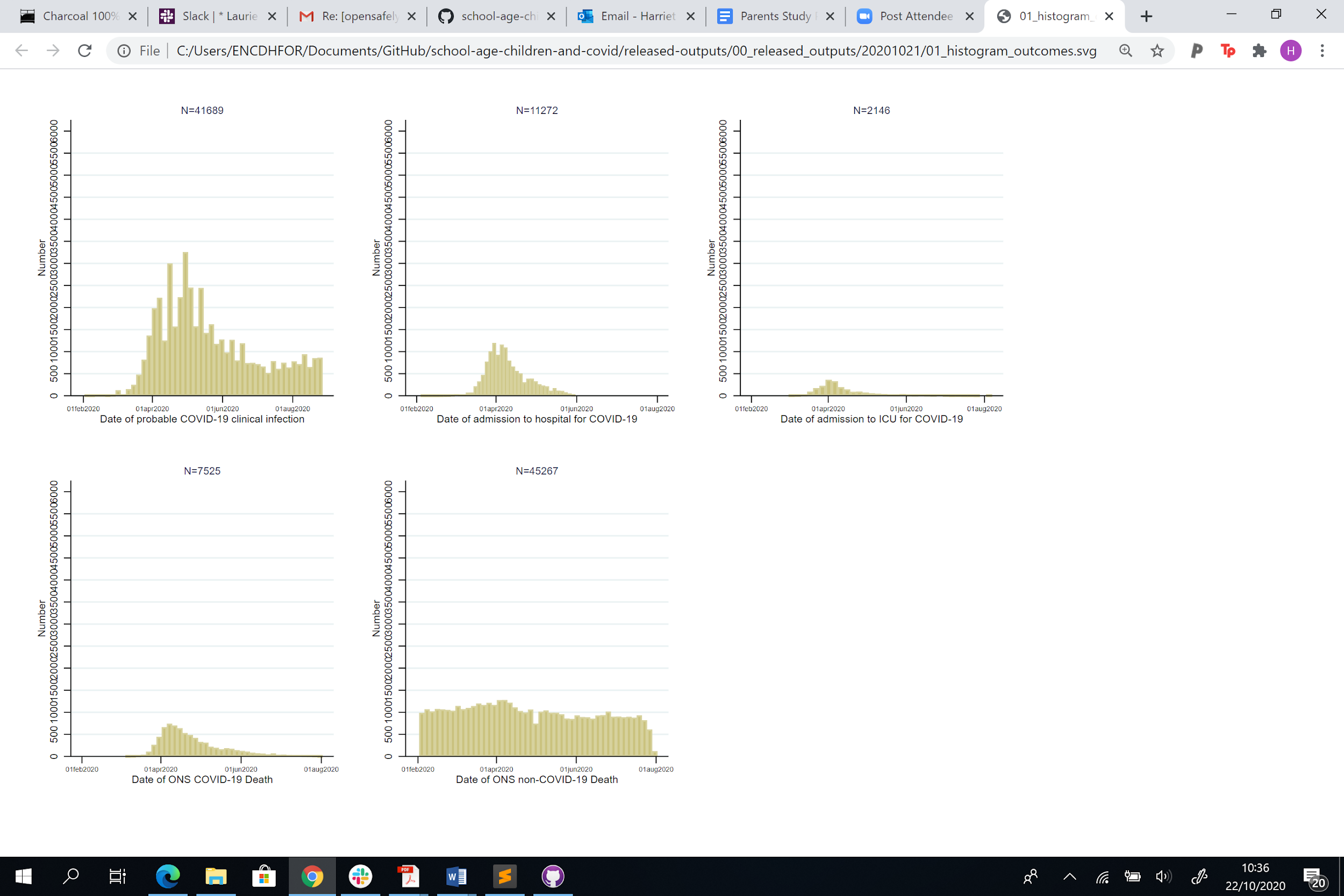

### **Figure A3: Results from sensitivity analysis: Hazard Ratios (HRs) for each COVID-19 outcome (a) evidence of SARS-CoV-2 infection recorded in primary care, (b) COVID-19 hospital admission, (c) COVID-19 ICU admission and (d) COVID-19 death), stratified by age**

**(a) Recorded SARS-CoV-2 infection**

**Adults 65 years and under**

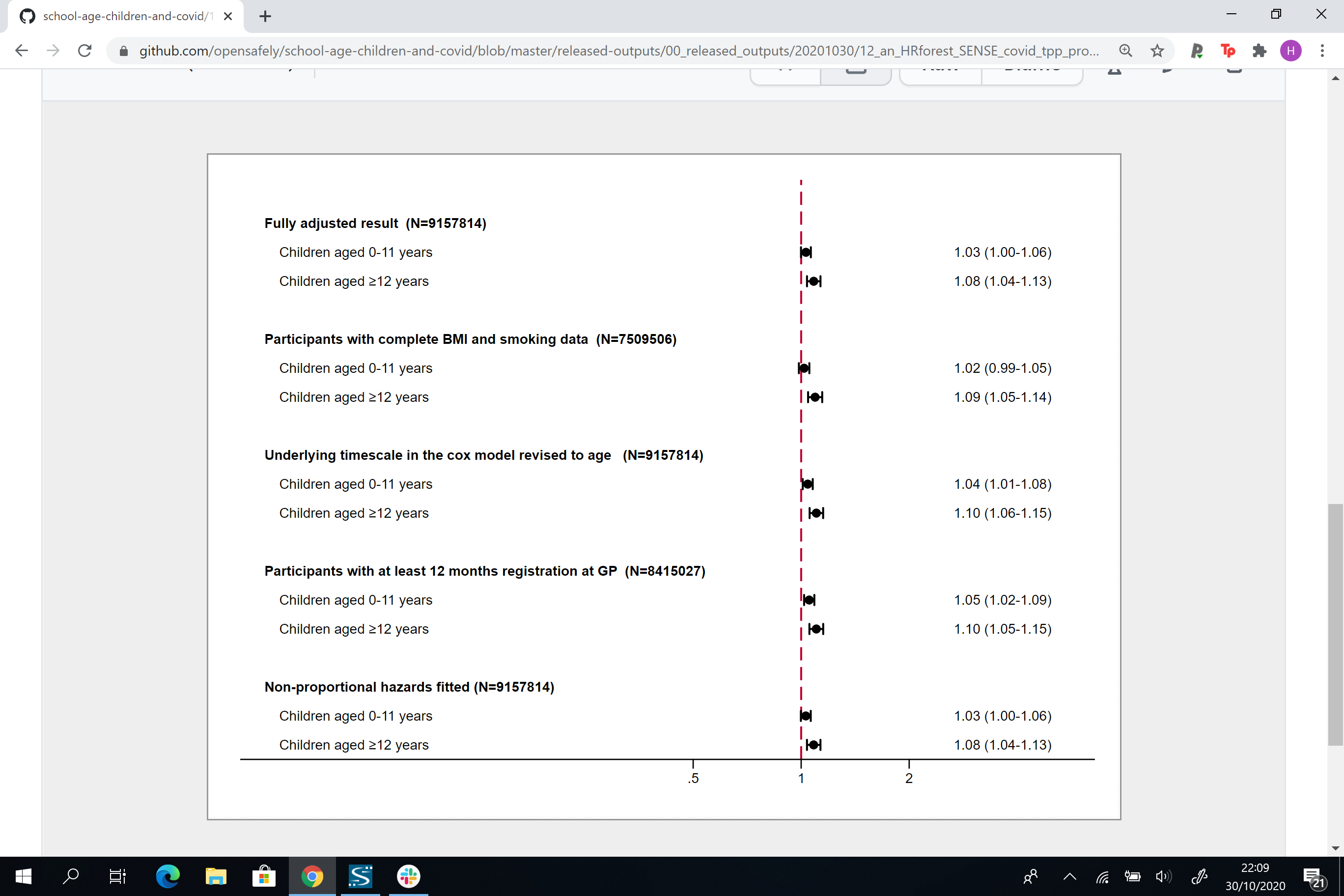

**Adults over 65 years**

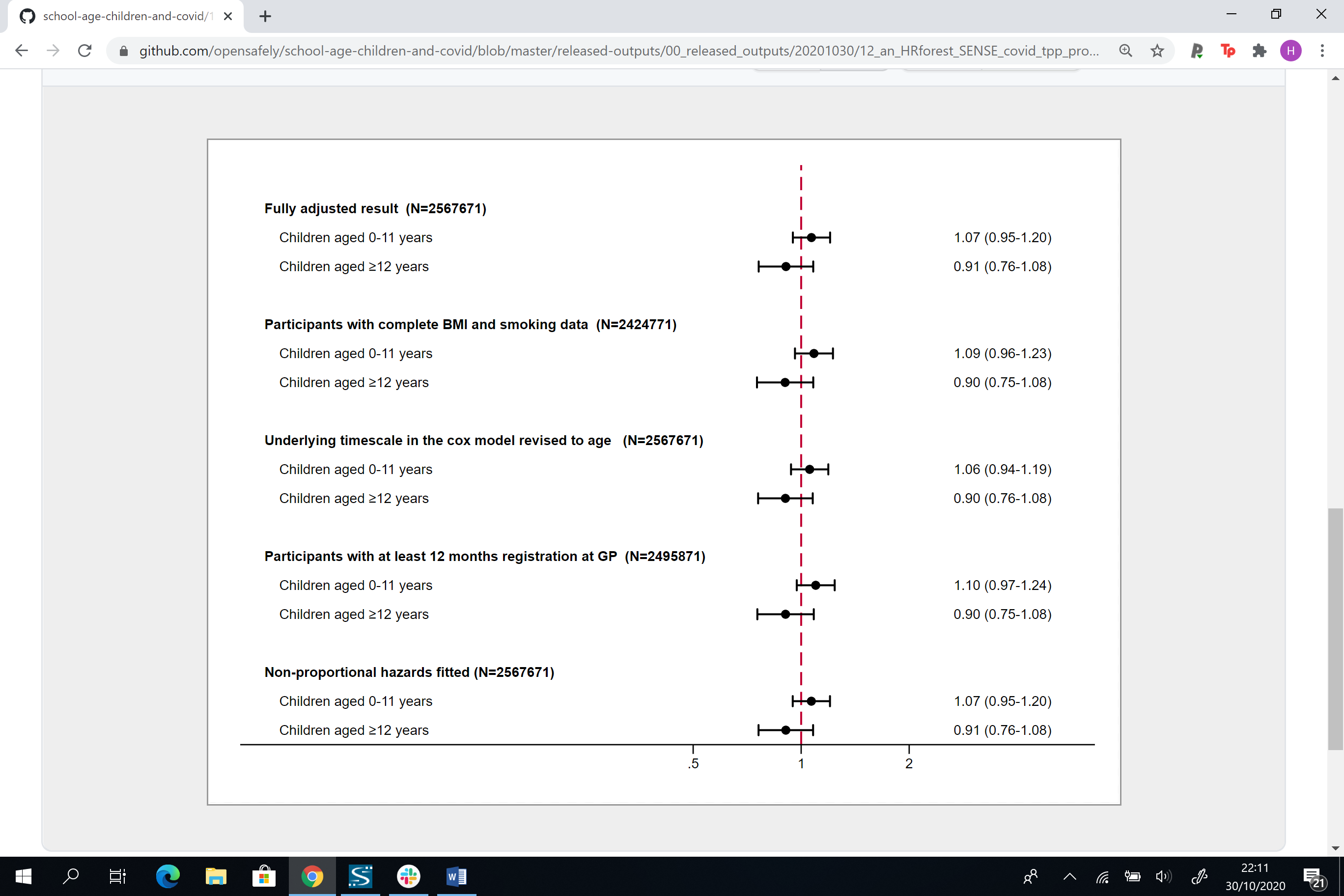

**(b) COVID-19 hospital admission**

**Adults 65 years and under**

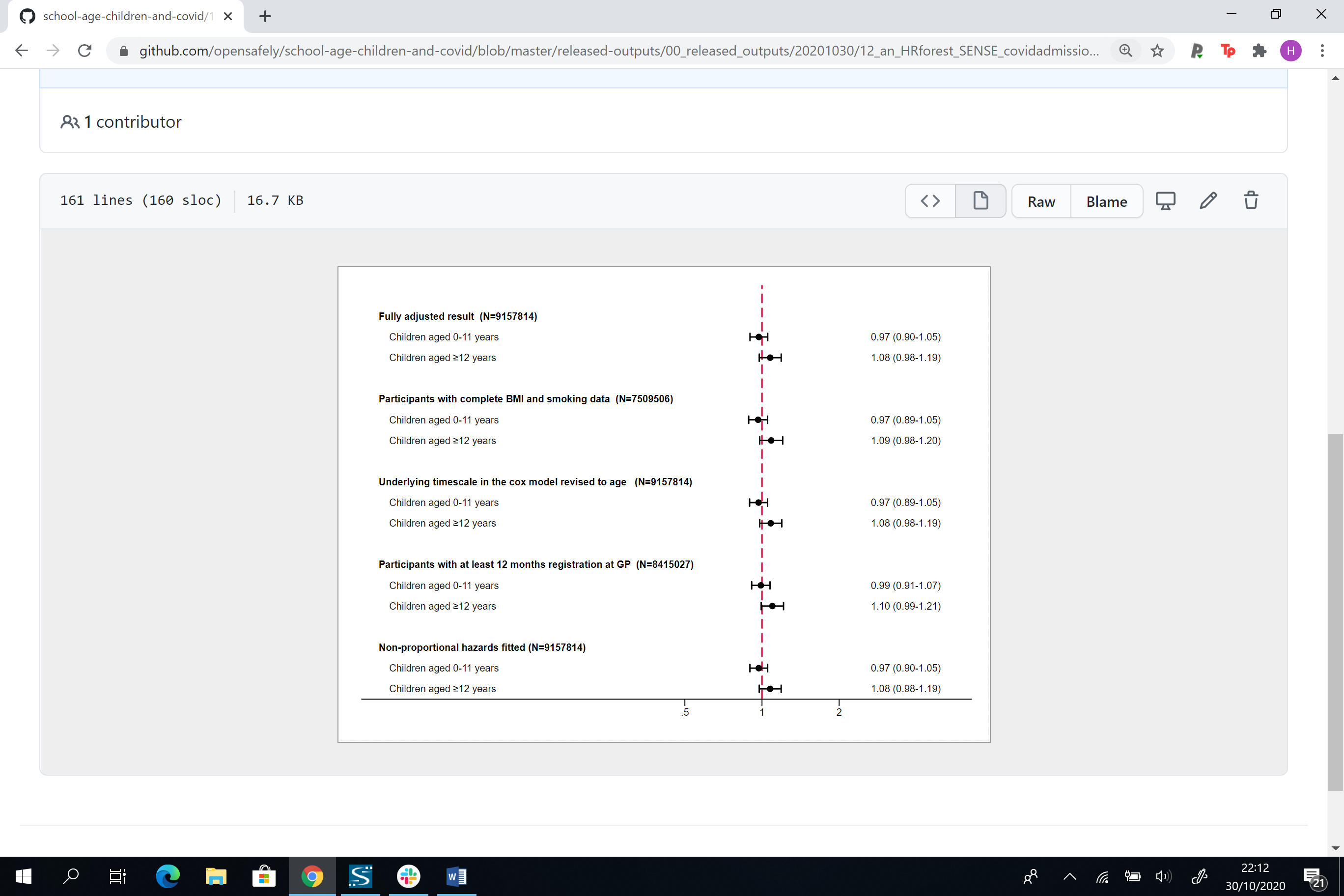

**Adults over 65 years**

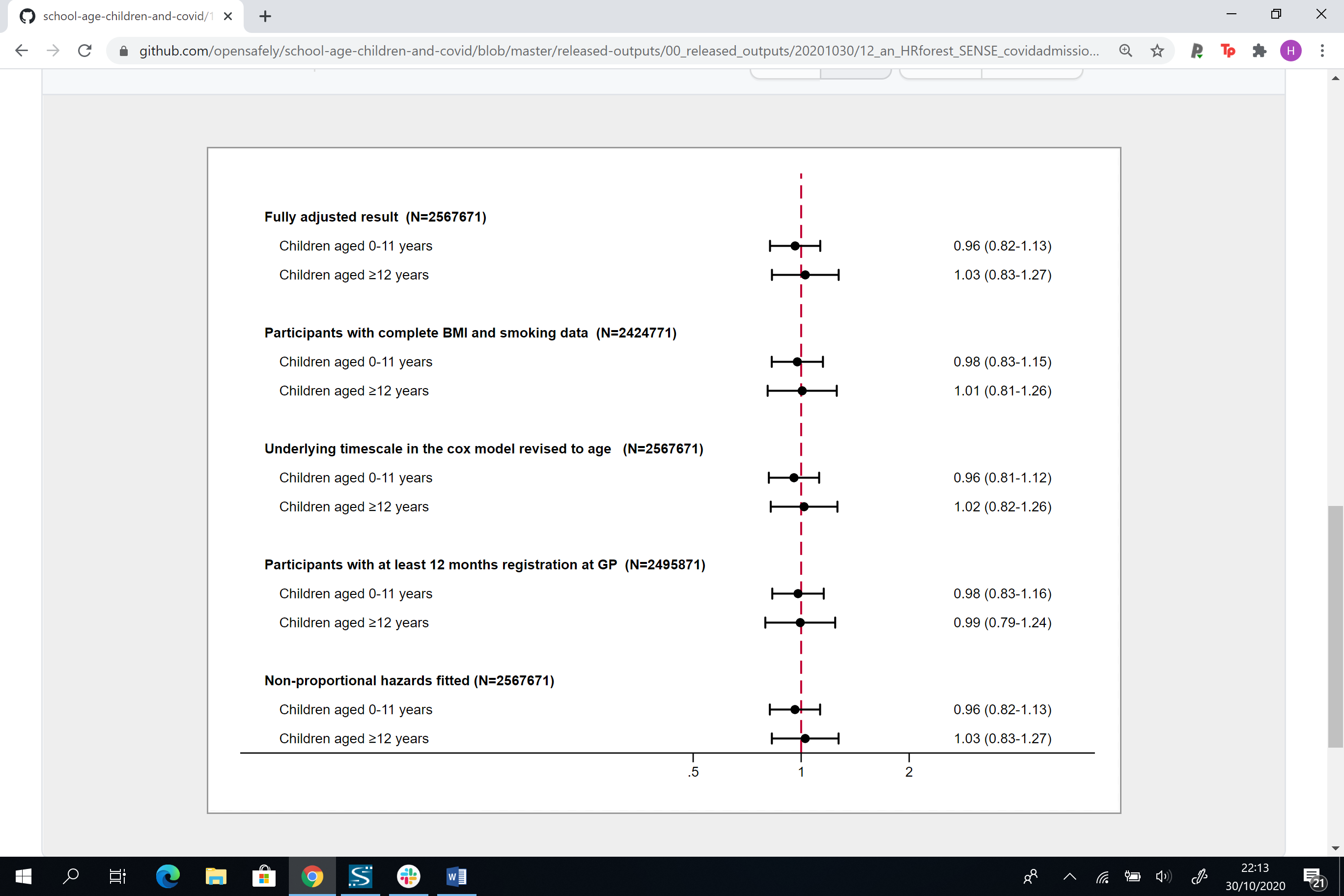

**(c) COVID-19 ICU admission**

**Adults 65 years and under**

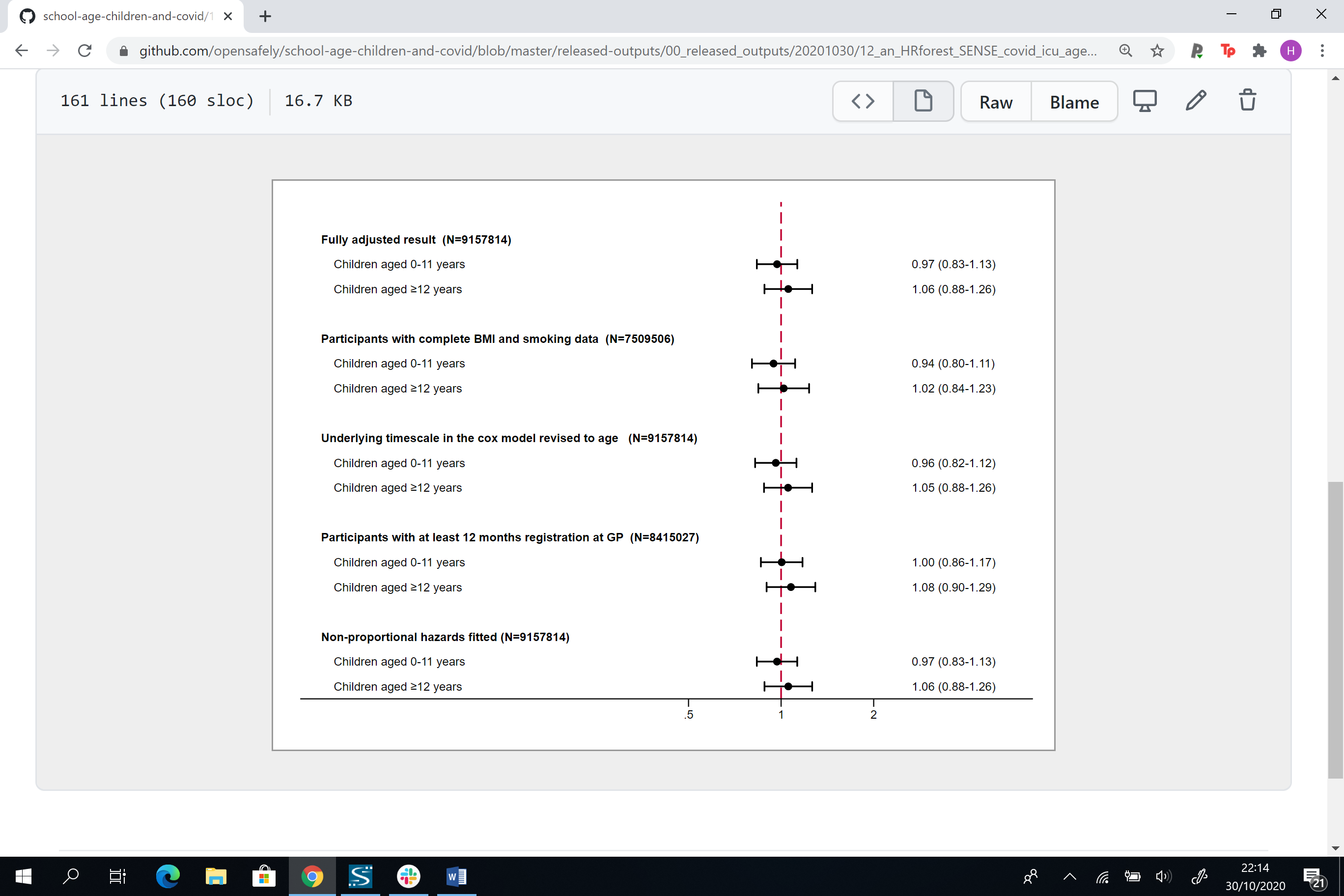

**Adults over 65 years**

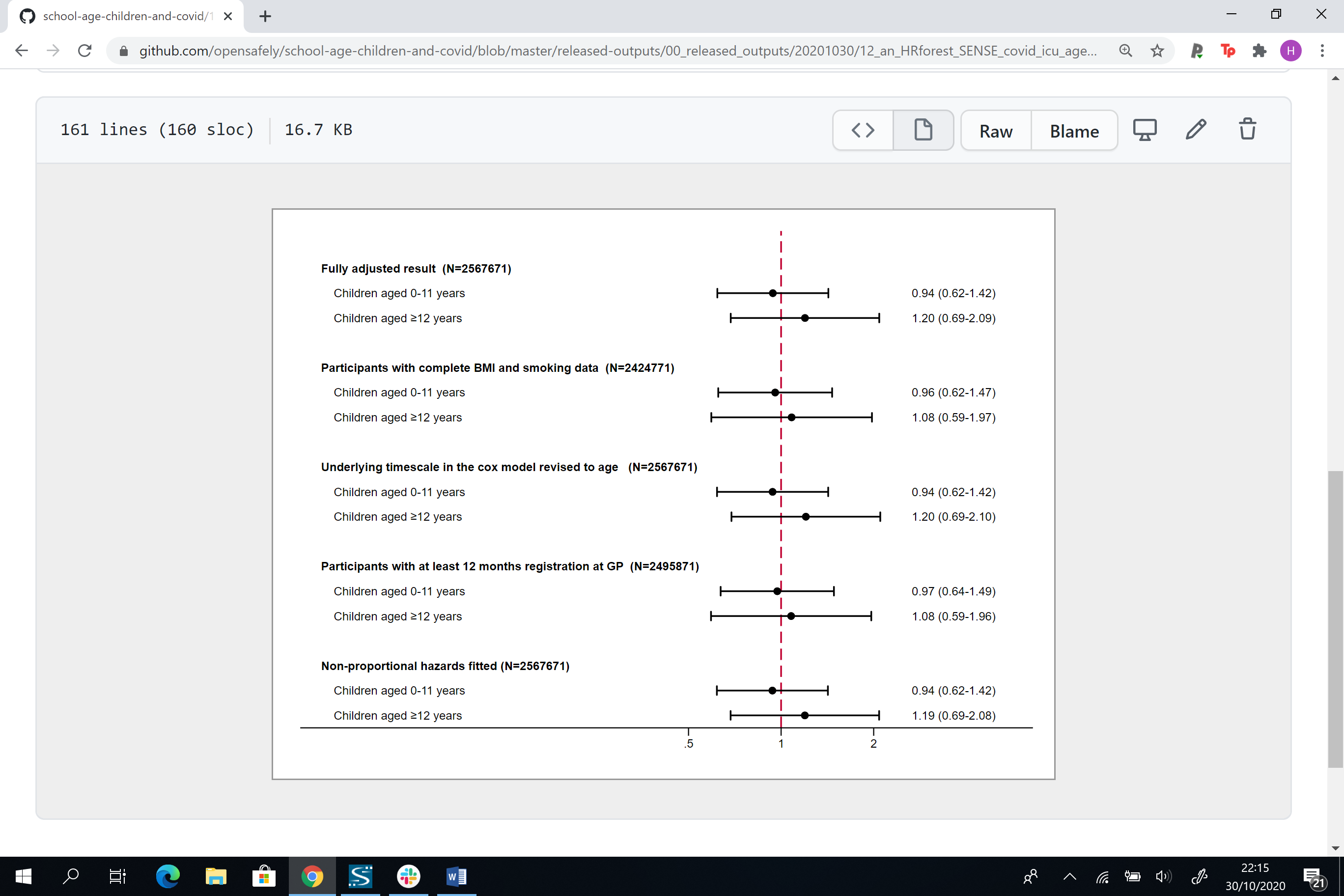

**(d) COVID-19 death**

**Adults 65 years and under**

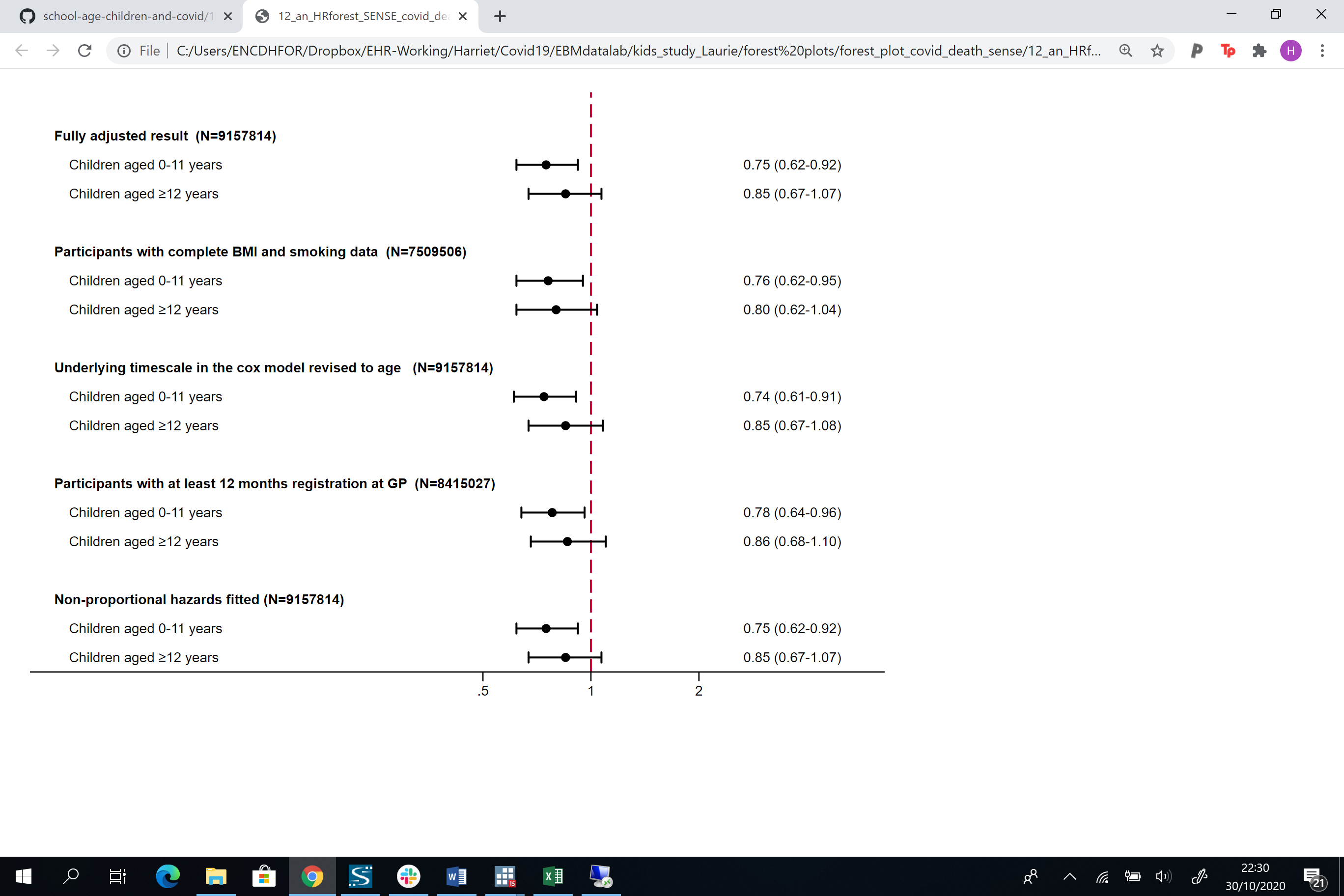

**Adults over 65 years**

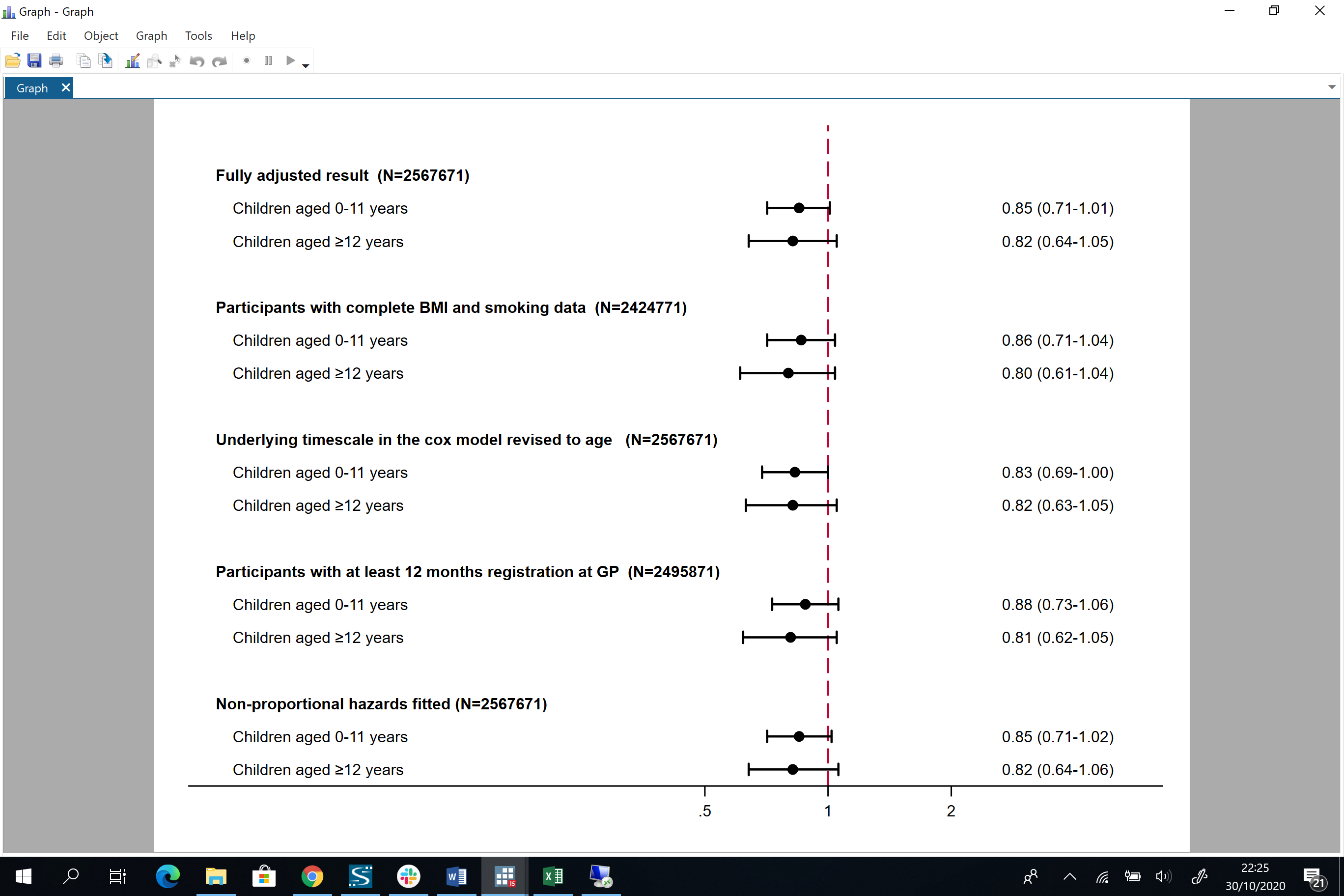

**Figure A4: Directed acyclic graph (DAG) illustrating implicitly assumed causal structure between household exposure to children and recorded SARS-CoV-2 infection, and severe outcomes from COVID-19**

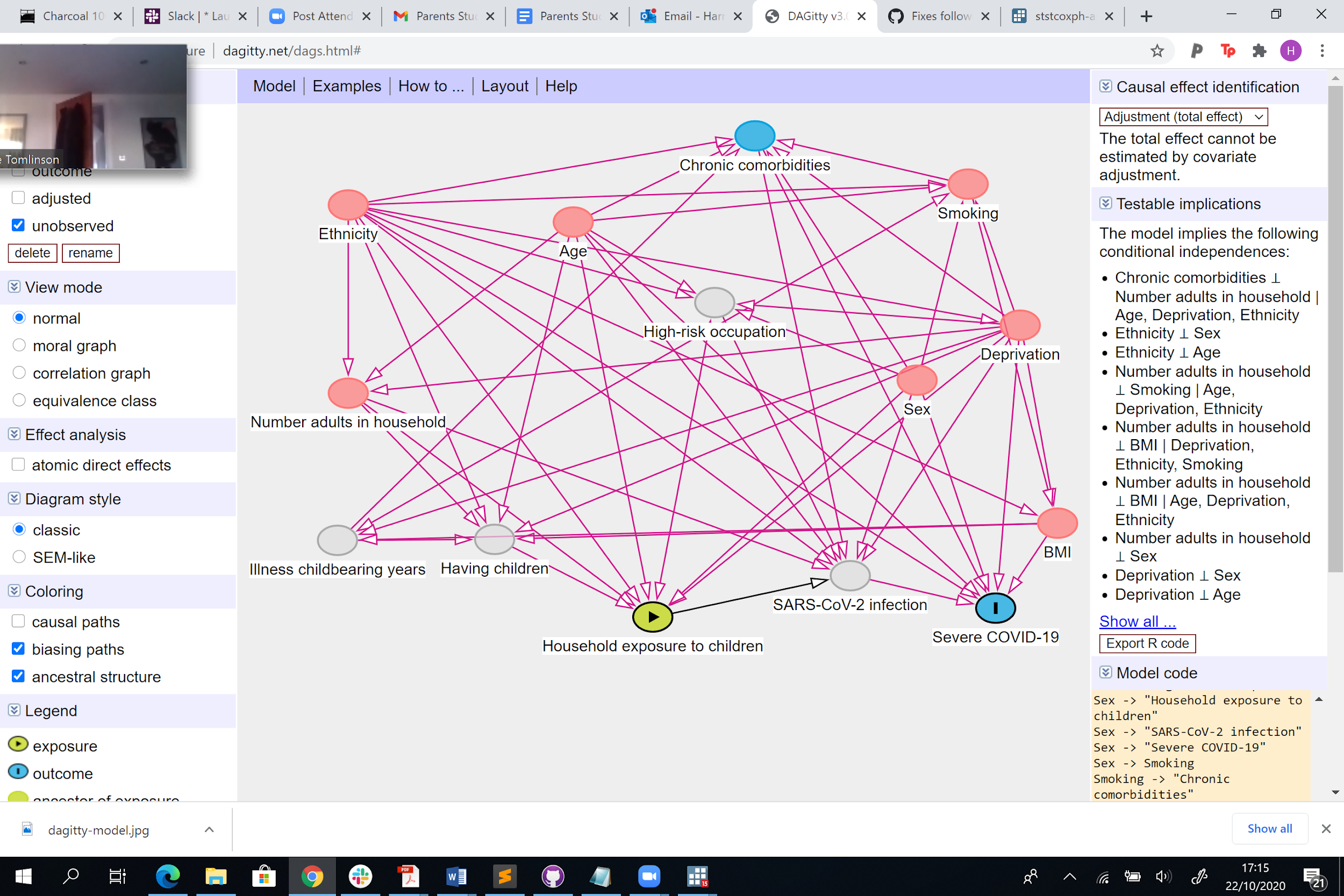

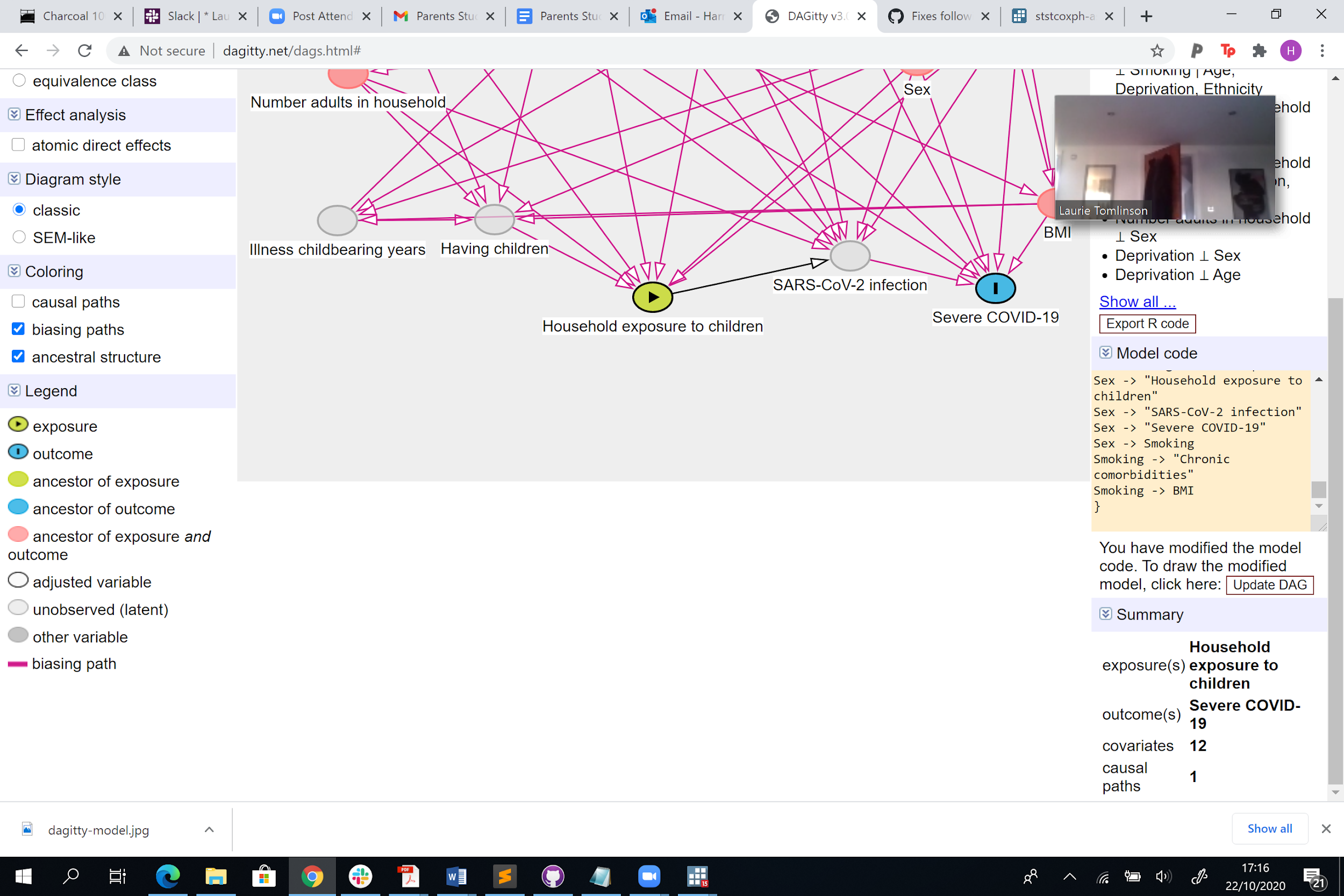

**Figure A5. Comorbidity adjusted hazard ratios (HRs) and 95% confidence intervals (CI) for each COVID-19 outcome, compared to having no children in the household by (a) sex, (b) time periods before and after 3rd April 2020 and (c) shielding status among those 65 years and over.**

a) **Sex**

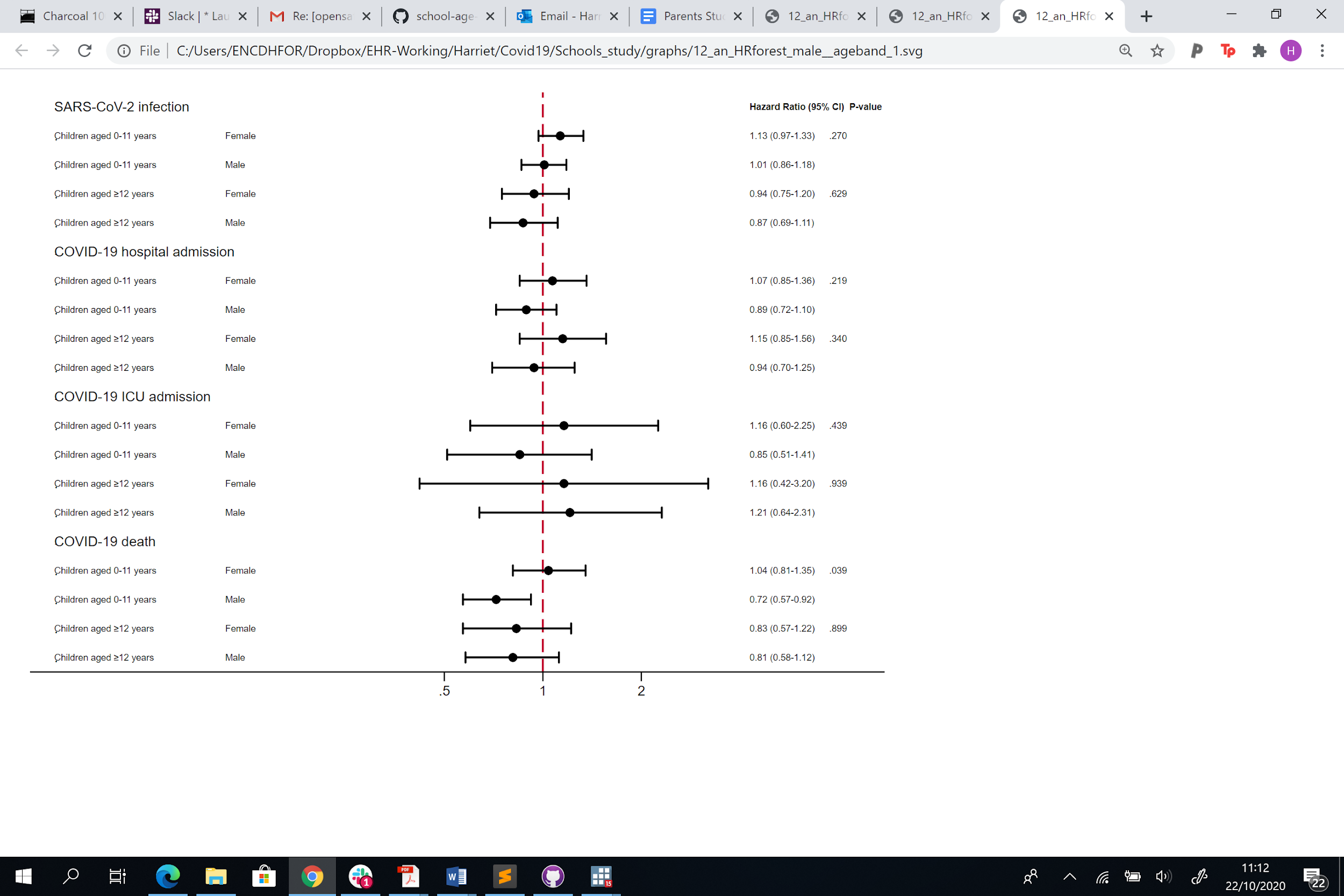

b) **Time periods before and on/after 3rd April 2020**

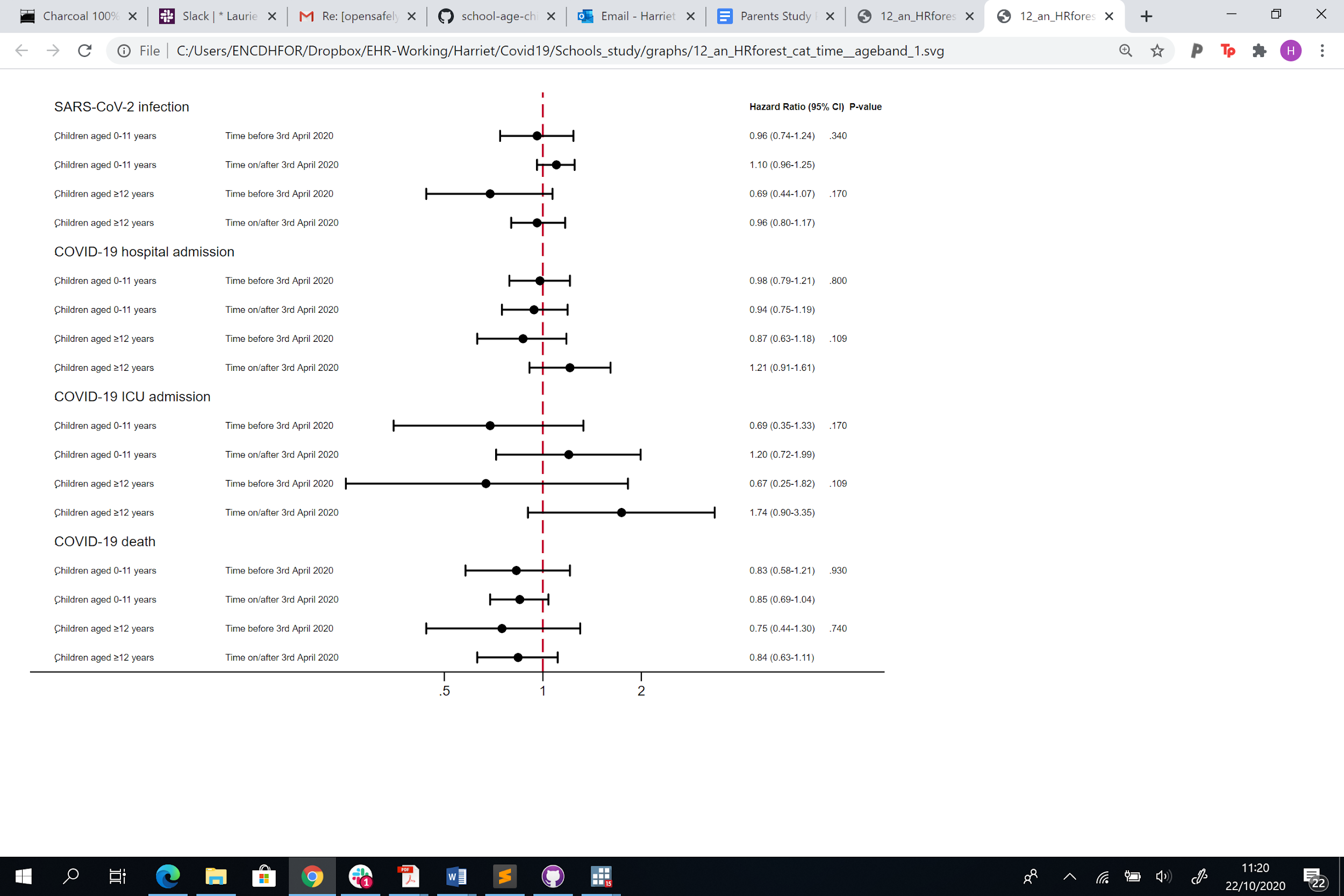

**c) Shielding status**

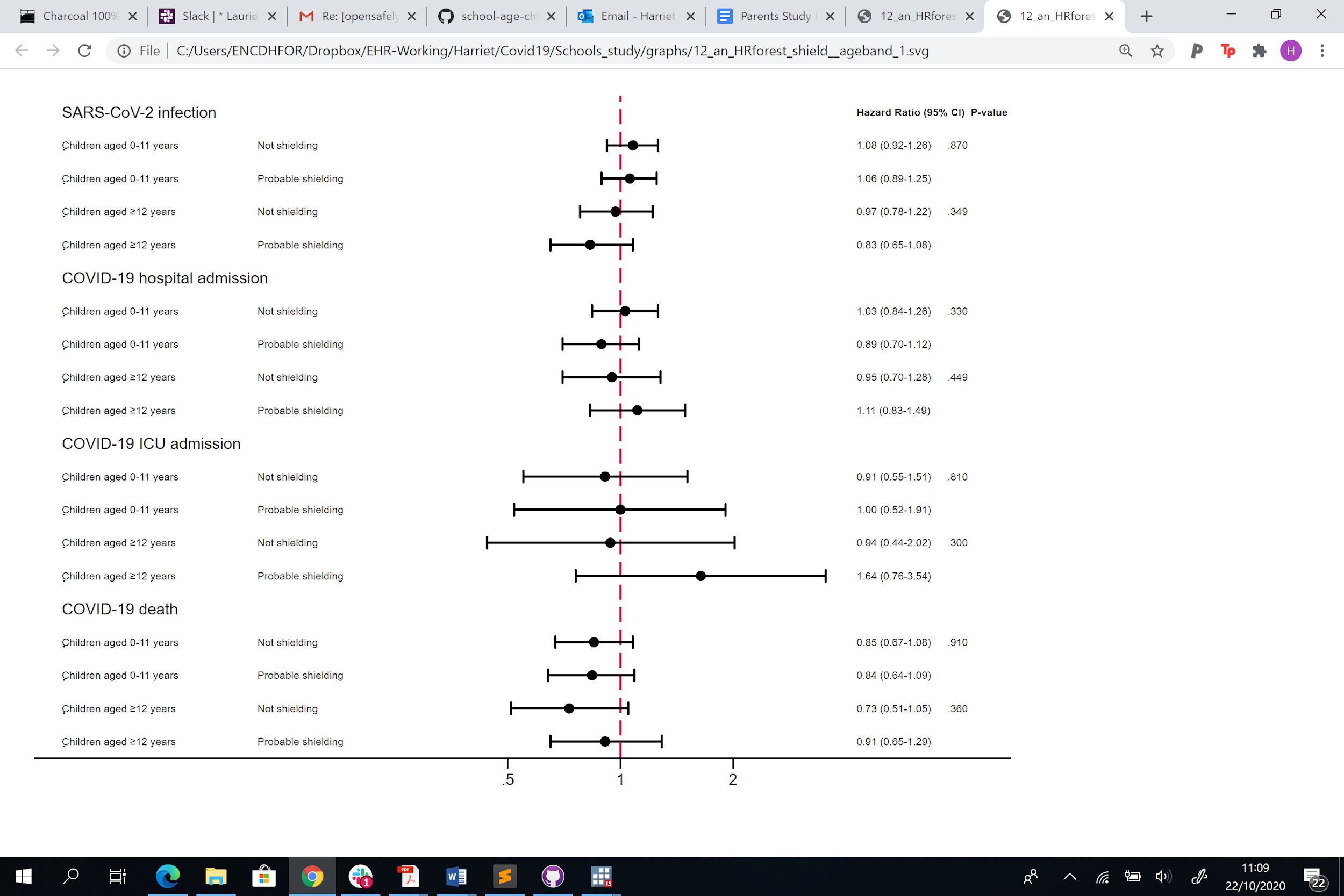

**Table A1: Changes to the original study protocol**

| **Suggestion** | **Rationale** |
| --- | --- |
| Stratify all results on age above and below 65 years. | We were concerned that our age adjustment was not sufficient to capture the variation in risk of infection with SARS-CoV-2 among those working and after retirement. |
| Separate COVID death and ICU admission | In early analyses the effect of living with children was in different directions for death from COVID-19 and ICU admission with COVID-19. In addition we had confirmed that there was adequate power to model each outcome separately. |
| Add non-COVID 19 death as an outcome | To contextualise our findings by comparing the risk of death from COVID-19 to and other causes. |
| Adjust for number of adults in household, rather than total number of people in the household | Data exploration showed that most of the additional people in a household were children, so results would have been hard to interpret if the ‘dose-response’ effect was also incorporated in the main model |
| Drop the planned sensitivity analysis using an alternative definition of COVID-19 in primary care to include diagnostic codes for suspected cases. | Exploration of suspected COVID-19 cases in primary care suggested there could be substantial misclassification of this outcome |
| Add outcome hospital admission for COVID-19 | This outcome became available during the time course of the study, and it enabled us to examine whether the findings were consistent with those for ICU admission |
| Re-run analysis with age as the underlying timescale, as a sensitivity analysis | To confirm we are adequately adjusting for age. |
| Developing a directed-acyclic graph to inform our model covariates | To confirm we are adjusting for all appropriate variables and not introducing collider bias. |
| Additional model fitting a time-interaction with variables where there was evidence of non-proportional hazards | To examine whether introducing time interactions where there was evidence of non-proportional hazards materially altered our findings. |
| Use multiple imputation to impute missing data for ethnicity | To examine whether the reduced study population introduced by adjusting for ethnicity altered our results in a meaningful way. |

**Table A2. Cohort description by outcomes (evidence of SARS-CoV-2 infection recorded in primary care, COVID-19 outcomes and non-COVID 19 deaths) for adults** $\leq$**65 years**

|  |  | **Number (%) within stratum)** | | | | |
| --- | --- | --- | --- | --- | --- | --- |
|  | **N (column %)** | **Recorded SARS-CoV-2 infection** | **COVID-19 hospital admissions** | **COVID-19 ICU admissions** | **COVID-19 deaths** | **Non-COVID-19 deaths** |
| **Total** | 9157814 (100.0) | 29863 (0.33) | 4776 (0.05) | 1471 (0.02) | 1173 (0.01) | 7788 (0.09) |
| **Age** |  |  |  |  |  |  |
| 18-<30 | 1947216 (21.3) | 4904 (0.25) | 246 (0.01) | 38 (0.00) | 13 (0.00) | 131 (0.01) |
| 30-<40 | 2202357 (24.0) | 6464 (0.29) | 552 (0.03) | 114 (0.01) | 43 (0.00) | 369 (0.02) |
| 40-<50 | 1991653 (21.7) | 6999 (0.35) | 1027 (0.05) | 295 (0.01) | 149 (0.01) | 1161 (0.06) |
| 50-<60 | 2012258 (22.0) | 7932 (0.39) | 1795 (0.09) | 602 (0.03) | 450 (0.02) | 3012 (0.15) |
| 60-<66 | 1004330 (11.0) | 3564 (0.35) | 1156 (0.12) | 422 (0.04) | 518 (0.05) | 3115 (0.31) |
| **Sex** |  |  |  |  |  |  |
| Female | 4714908 (51.5) | 17285 (0.37) | 1977 (0.04) | 466 (0.01) | 414 (0.01) | 3252 (0.07) |
| Male | 4442906 (48.5) | 12578 (0.28) | 2799 (0.06) | 1005 (0.02) | 759 (0.02) | 4536 (0.10) |
| **BMI (kg/m2)** |  |  |  |  |  |  |
| <18.5 | 178486 (1.9) | 458 (0.26) | 51 (0.03) | 9 (0.01) | 21 (0.01) | 430 (0.24) |
| 18.5-24.9 | 2843580 (31.1) | 7466 (0.26) | 705 (0.02) | 166 (0.01) | 184 (0.01) | 2314 (0.08) |
| 25-29.9 | 2497554 (27.3) | 8740 (0.35) | 1368 (0.05) | 395 (0.02) | 309 (0.01) | 1987 (0.08) |
| 30-34.9 (Obese class I) | 1252559 (13.7) | 5441 (0.43) | 1127 (0.09) | 385 (0.03) | 241 (0.02) | 1197 (0.10) |
| 35-39.9 (Obese class II) | 505136 (5.5) | 2537 (0.50) | 633 (0.13) | 214 (0.04) | 172 (0.03) | 565 (0.11) |
| ≥40 (Obese class III) | 270158 (3.0) | 1677 (0.62) | 436 (0.16) | 158 (0.06) | 120 (0.04) | 448 (0.17) |
| *Missing* | 1610341 (17.6) | 3544 (0.22) | 456 (0.03) | 144 (0.01) | 126 (0.01) | 847 (0.05) |
| **Smoking** |  |  |  |  |  |  |
| Never | 4405852 (48.1) | 15902 (0.36) | 2513 (0.06) | 743 (0.02) | 491 (0.01) | 2096 (0.05) |
| Former | 2636324 (28.8) | 9605 (0.36) | 1855 (0.07) | 623 (0.02) | 496 (0.02) | 2768 (0.10) |
| Current | 1869068 (20.4) | 3932 (0.21) | 382 (0.02) | 98 (0.01) | 181 (0.01) | 2886 (0.15) |
| *Missing* | 246570 (2.7) | 424 (0.17) | 26 (0.01) | 7 (0.00) | REDACTED | 38 (0.02) |
| **Ethnicity** |  |  |  |  |  |  |
| White | 7569454 (82.7) | 20132 (0.27) | 3078 (0.04) | 925 (0.01) | 795 (0.01) | 7046 (0.09) |
| Mixed | 158349 (1.7) | 566 (0.36) | 126 (0.08) | 44 (0.03) | 28 (0.02) | 59 (0.04) |
| South Asian | 832720 (9.1) | 6794 (0.82) | 897 (0.11) | 293 (0.04) | 211 (0.03) | 389 (0.05) |
| Black | 306539 (3.3) | 1527 (0.50) | 458 (0.15) | 140 (0.05) | 99 (0.03) | 213 (0.07) |
| Other | 290752 (3.2) | 844 (0.29) | 217 (0.07) | 69 (0.02) | 40 (0.01) | 81 (0.03) |
| **IMD quintile** |  |  |  |  |  |  |
| 1 (least deprived) | 1684917 (18.4) | 4108 (0.24) | 680 (0.04) | 185 (0.01) | 118 (0.01) | 1017 (0.06) |
| 2 | 1805617 (19.7) | 4968 (0.28) | 791 (0.04) | 233 (0.01) | 169 (0.01) | 1230 (0.07) |
| 3 | 1874964 (20.5) | 5547 (0.30) | 916 (0.05) | 283 (0.02) | 214 (0.01) | 1460 (0.08) |
| 4 | 1976632 (21.6) | 7117 (0.36) | 1160 (0.06) | 380 (0.02) | 337 (0.02) | 1782 (0.09) |
| 5 (most deprived) | 1815684 (19.8) | 8123 (0.45) | 1229 (0.07) | 390 (0.02) | 335 (0.02) | 2299 (0.13) |
| **Total number adults in household** |  |  |  |  |  |  |
| 1 | 2226830 (24.3) | 6017 (0.27) | 1112 (0.05) | 320 (0.01) | 326 (0.01) | 2764 (0.12) |
| 2 | 3779868 (41.3) | 11743 (0.31) | 1773 (0.05) | 522 (0.01) | 376 (0.01) | 2922 (0.08) |
| ⋝3 | 3151116 (34.4) | 12103 (0.38) | 1891 (0.06) | 629 (0.02) | 471 (0.01) | 2102 (0.07) |
| **Blood pressure** |  |  |  |  |  |  |
| Normal | 2440215 (26.6) | 7720 (0.32) | 934 (0.04) | 197 (0.01) | 227 (0.01) | 1875 (0.08) |
| Elevated | 1324098 (14.5) | 4330 (0.33) | 576 (0.04) | 167 (0.01) | 141 (0.01) | 1011 (0.08) |
| High Stage 1 | 2878905 (31.4) | 10371 (0.36) | 1831 (0.06) | 596 (0.02) | 435 (0.02) | 2485 (0.09) |
| High Stage 2 | 1549272 (16.9) | 5960 (0.38) | 1335 (0.09) | 477 (0.03) | 355 (0.02) | 2296 (0.15) |
| Missing | 965324 (10.5) | 1482 (0.15) | 100 (0.01) | 34 (0.00) | 15 (0.00) | 121 (0.01) |
| **High bp or diagnosed hypertension** | 2156211 (23.5) | 9205 (0.43) | 2313 (0.11) | 835 (0.04) | 709 (0.03) | 3957 (0.18) |
| ***Comorbidities*** |  |  |  |  |  |  |
| **Chronic respiratory disease ex asthma** | 184872 (2.0) | 1150 (0.62) | 388 (0.21) | 127 (0.07) | 173 (0.09) | 1398 (0.76) |
| **Asthma** | 1574912 (17.2) | 5887 (0.37) | 1057 (0.07) | 305 (0.02) | 234 (0.01) | 1452 (0.09) |
| **Chronic cardiac disease** | 259057 (2.8) | 1544 (0.60) | 563 (0.22) | 165 (0.06) | 233 (0.09) | 1422 (0.55) |
| **Diabetes** |  |  |  |  |  |  |
| No diabetes | 8712703 (95.1) | 26661 (0.31) | 3665 (0.04) | 1039 (0.01) | 761 (0.01) | 6152 (0.07) |
| Type 1, controlled | 12483 (0.1) | 50 (0.40) | 17 (0.14) | REDACTED | 6 (0.05) | 46 (0.37) |
| Type 1, uncontrolled | 37382 (0.4) | 193 (0.52) | 55 (0.15) | 19 (0.05) | 20 (0.05) | 140 (0.37) |
| Type 2, controlled | 219161 (2.4) | 1509 (0.69) | 474 (0.22) | 182 (0.08) | 172 (0.08) | 743 (0.34) |
| Type 2, uncontrolled | 171932 (1.9) | 1428 (0.83) | 556 (0.32) | 221 (0.13) | 208 (0.12) | 686 (0.40) |
| Diabetes, no HbA1c | 4153 (0.0) | 22 (0.53) | 9 (0.22) | REDACTED | 6 (0.14) | 21 (0.51) |
| **Haematological cancer** |  |  |  |  |  |  |
| Diagnosed < 1 year ago | 2349 (0.0) | 31 (1.32) | 9 (0.38) | REDACTED | 7 (0.30) | 66 (2.81) |
| Diagnosed 1-4.9 years ago | 7332 (0.1) | 66 (0.90) | 35 (0.48) | 9 (0.12) | 14 (0.19) | 97 (1.32) |
| Diagnosed ≥5 years ago | 19225 (0.2) | 105 (0.55) | 24 (0.12) | 14 (0.07) | 13 (0.07) | 81 (0.42) |
| **Non-haematological**  **cancer** |  |  |  |  |  |  |
| Diagnosed < 1 year ago | 22171 (0.2) | 150 (0.68) | 48 (0.22) | 7 (0.03) | 33 (0.15) | 1053 (4.75) |
| Diagnosed 1-4.9 years ago | 64176 (0.7) | 269 (0.42) | 68 (0.11) | 17 (0.03) | 46 (0.07) | 1107 (1.72) |
| Diagnosed ≥5 years ago | 113243 (1.2) | 433 (0.38) | 99 (0.09) | 18 (0.02) | 47 (0.04) | 606 (0.54) |
| **Reduced kidney function** |  |  |  |  |  |  |
| Estimated GFR 30-60 | 93017 (1.0) | 539 (0.58) | 225 (0.24) | 85 (0.09) | 127 (0.14) | 545 (0.59) |
| Estimated GFR <30 | 9067 (0.1) | 194 (2.14) | 114 (1.26) | 28 (0.31) | 63 (0.69) | 261 (2.88) |
| **End-stage renal disease*** | 10815 (0.1) | 225 (2.08) | 125 (1.16) | 41 (0.38) | 49 (0.45) | 164 (1.52) |
| **Chronic Liver disease** | 51404 (0.6) | 327 (0.64) | 105 (0.20) | 23 (0.04) | 58 (0.11) | 765 (1.49) |
| **Stroke/dementia** | 71425 (0.8) | 464 (0.65) | 169 (0.24) | 60 (0.08) | 88 (0.12) | 566 (0.79) |
| **Other neurological disease** | 63572 (0.7) | 299 (0.47) | 120 (0.19) | 18 (0.03) | 63 (0.10) | 315 (0.50) |
| **Solid organ transplant**** | 2103 (0.0) | 31 (1.47) | 10 (0.48) | REDACTED | REDACTED | 21 (1.00) |
| **Asplenia** | 10867 (0.1) | 84 (0.77) | 16 (0.15) | 9 (0.08) | REDACTED | 69 (0.63) |
| **Rheumatoid/Lupus/ Psoriasis** | 409631 (4.5) | 1653 (0.40) | 334 (0.08) | 109 (0.03) | 98 (0.02) | 614 (0.15) |
| **Other immunosuppressive condition** | 32745 (0.4) | 242 (0.74) | 41 (0.13) | 16 (0.05) | 21 (0.06) | 115 (0.35) |
| **Shielding***** | 1922340 (21.0) | 7836 (0.41) | 1648 (0.09) | 494 (0.03) | 518 (0.03) | 5136 (0.27) |

*End-stage renal disease includes on dialysis or having had a kidney transplant

**All solid organ transplants, excluding kidney

***Shielding includes organ transplant recipients, renal replacement therapy, haematological cancers, non-haematological cancers, immunodeficiencies/asplenia and severe respiratory conditions.

**Table A3. Cohort description by outcomes (evidence of SARS-CoV-2 infection recorded in primary care, COVID-19 outcomes and non-COVID 19 deaths) for adults** $\boldsymbol{>}$**65 years**

|  |  | **Number (%) within stratum)** | | | | |
| --- | --- | --- | --- | --- | --- | --- |
|  | **N (column %)** | **Recorded SARS-CoV-2 infection** | **COVID-19 hospital admissions** | **COVID-19 ICU admissions** | **COVID-19 deaths** | **Non- COVID-19 deaths** |
| **Total** | 2567671 (100.0) | 11826 (0.46) | 6496 (0.25) | 675 (0.03) | 6352 (0.25) | 37479 (1.46) |
| **Age** |  |  |  |  |  |  |
| >65-70 | 609634 (23.7) | 1856 (0.30) | 778 (0.13) | 222 (0.04) | 453 (0.07) | 2921 (0.48) |
| 70-80 | 1300326 (50.6) | 4612 (0.35) | 2480 (0.19) | 395 (0.03) | 1975 (0.15) | 12093 (0.93) |
| 80+ | 657711 (25.6) | 5358 (0.81) | 3238 (0.49) | 58 (0.01) | 3924 (0.60) | 22465 (3.42) |
| **Sex** |  |  |  |  |  |  |
| Female | 1384514 (53.9) | 5774 (0.42) | 2672 (0.19) | 189 (0.01) | 2635 (0.19) | 18130 (1.31) |
| Male | 1183157 (46.1) | 6052 (0.51) | 3824 (0.32) | 486 (0.04) | 3717 (0.31) | 19349 (1.64) |
| **BMI (kg/m2)** |  |  |  |  |  |  |
| <18.5 | 44163 (1.7) | 439 (0.99) | 176 (0.40) | REDACTED | 251 (0.57) | 2439 (5.52) |
| 18.5-24.9 | 760180 (29.6) | 3506 (0.46) | 1731 (0.23) | 109 (0.01) | 1956 (0.26) | 14001 (1.84) |
| 25-29.9 | 949715 (37.0) | 3856 (0.41) | 2139 (0.23) | 257 (0.03) | 1896 (0.20) | 10712 (1.13) |
| 30-34.9 (Obese class I) | 462119 (18.0) | 2257 (0.49) | 1346 (0.29) | 173 (0.04) | 1163 (0.25) | 4980 (1.08) |
| 35-39.9 (Obese class II) | 151009 (5.9) | 859 (0.57) | 554 (0.37) | 78 (0.05) | 474 (0.31) | 1847 (1.22) |
| ≥40 (Obese class III) | 58570 (2.3) | 403 (0.69) | 237 (0.40) | 24 (0.04) | 214 (0.37) | 823 (1.41) |
| *Missing* | 141915 (5.5) | 506 (0.36) | 313 (0.22) | 31 (0.02) | 398 (0.28) | 2677 (1.89) |
| **Smoking** |  |  |  |  |  |  |
| Never | 1011539 (39.4) | 3901 (0.39) | 2005 (0.20) | 203 (0.02) | 1871 (0.18) | 11149 (1.10) |
| Former | 1336686 (52.1) | 7063 (0.53) | 4097 (0.31) | 443 (0.03) | 4020 (0.30) | 21924 (1.64) |
| Current | 213854 (8.3) | 843 (0.39) | 386 (0.18) | 27 (0.01) | 450 (0.21) | 4344 (2.03) |
| *Missing* | 5592 (0.2) | 19 (0.34) | 8 (0.14) | REDACTED | 11 (0.20) | 62 (1.11) |
| **Ethnicity** |  |  |  |  |  |  |
| White | 2419165 (94.2) | 10622 (0.44) | 5631 (0.23) | 558 (0.02) | 5664 (0.23) | 35648 (1.47) |
| Mixed | 9797 (0.4) | 54 (0.55) | 44 (0.45) | 7 (0.07) | 34 (0.35) | 116 (1.18) |
| South Asian | 90017 (3.5) | 809 (0.90) | 513 (0.57) | 64 (0.07) | 453 (0.50) | 1159 (1.29) |
| Black | 27290 (1.1) | 226 (0.83) | 220 (0.81) | 28 (0.10) | 144 (0.53) | 363 (1.33) |
| Other | 21402 (0.8) | 115 (0.54) | 88 (0.41) | 18 (0.08) | 57 (0.27) | 193 (0.90) |
| **IMD quintile** |  |  |  |  |  |  |
| 1 (least deprived) | 618192 (24.1) | 2134 (0.35) | 1066 (0.17) | 135 (0.02) | 1090 (0.18) | 7261 (1.17) |
| 2 | 596748 (23.2) | 2373 (0.40) | 1251 (0.21) | 132 (0.02) | 1263 (0.21) | 8013 (1.34) |
| 3 | 548147 (21.3) | 2388 (0.44) | 1320 (0.24) | 148 (0.03) | 1267 (0.23) | 7884 (1.44) |
| 4 | 460588 (17.9) | 2572 (0.56) | 1466 (0.32) | 146 (0.03) | 1386 (0.30) | 7515 (1.63) |
| 5 (most deprived) | 343996 (13.4) | 2359 (0.69) | 1393 (0.40) | 114 (0.03) | 1346 (0.39) | 6806 (1.98) |
| **Total number adults in household** |  |  |  |  |  |  |
| 1 | 877922 (34.2) | 5015 (0.57) | 2794 (0.32) | 170 (0.02) | 2978 (0.34) | 17344 (1.98) |
| 2 | 1319765 (51.4) | 4829 (0.37) | 2610 (0.20) | 358 (0.03) | 2383 (0.18) | 15286 (1.16) |
| ⋝3 | 369984 (14.4) | 1982 (0.54) | 1092 (0.30) | 147 (0.04) | 991 (0.27) | 4849 (1.31) |
| **Blood pressure** |  |  |  |  |  |  |
| Normal | 298019 (11.6) | 2043 (0.69) | 1144 (0.38) | 75 (0.03) | 1307 (0.44) | 8818 (2.96) |
| Elevated | 400138 (15.6) | 1955 (0.49) | 1143 (0.29) | 109 (0.03) | 1146 (0.29) | 6318 (1.58) |
| High Stage 1 | 947144 (36.9) | 3902 (0.41) | 2124 (0.22) | 246 (0.03) | 1929 (0.20) | 10998 (1.16) |
| High Stage 2 | 909480 (35.4) | 3910 (0.43) | 2075 (0.23) | 241 (0.03) | 1959 (0.22) | 11274 (1.24) |
| Missing | 12890 (0.5) | 16 (0.12) | 10 (0.08) | REDACTED | 11 (0.09) | 71 (0.55) |
| **High bp or diagnosed hypertension** | 1769252 (68.9) | 9002 (0.51) | 5045 (0.29) | 499 (0.03) | 4992 (0.28) | 28802 (1.63) |
| ***Comorbidities*** |  |  |  |  |  |  |
| **Chronic respiratory disease ex asthma** | 317732 (12.4) | 2956 (0.93) | 1682 (0.53) | 108 (0.03) | 1612 (0.51) | 9939 (3.13) |
| **Asthma** | 347416 (13.5) | 2119 (0.61) | 1115 (0.32) | 107 (0.03) | 969 (0.28) | 5063 (1.46) |
| **Chronic cardiac disease** | 536239 (20.9) | 4470 (0.83) | 2545 (0.47) | 153 (0.03) | 2739 (0.51) | 16020 (2.99) |
| **Diabetes** |  |  |  |  |  |  |
| No diabetes | 2103299 (81.9) | 8092 (0.38) | 4154 (0.20) | 472 (0.02) | 4070 (0.19) | 26858 (1.28) |
| Type 1, controlled | 3270 (0.1) | 29 (0.89) | 19 (0.58) | REDACTED | 18 (0.55) | 99 (3.03) |
| Type 1, uncontrolled | 6772 (0.3) | 75 (1.11) | 36 (0.53) | REDACTED | 45 (0.66) | 221 (3.26) |
| Type 2, controlled | 311147 (12.1) | 2306 (0.74) | 1381 (0.44) | 115 (0.04) | 1395 (0.45) | 6960 (2.24) |
| Type 2, uncontrolled | 141345 (5.5) | 1308 (0.93) | 893 (0.63) | 84 (0.06) | 813 (0.58) | 3287 (2.33) |
| Diabetes, no HbA1c | 1838 (0.1) | 16 (0.87) | 13 (0.71) | 0 (0.00) | 11 (0.60) | 54 (2.94) |
| **Haematological cancer** |  |  |  |  |  |  |
| Diagnosed < 1 year ago | 3661 (0.1) | 52 (1.42) | 28 (0.76) | REDACTED | 30 (0.82) | 286 (7.81) |
| Diagnosed 1-4.9 years ago | 11073 (0.4) | 116 (1.05) | 81 (0.73) | 13 (0.12) | 80 (0.72) | 474 (4.28) |
| Diagnosed ≥5 years ago | 22672 (0.9) | 204 (0.90) | 104 (0.46) | 8 (0.04) | 106 (0.47) | 713 (3.14) |
| **Non-haematological**  **cancer** |  |  |  |  |  |  |
| Diagnosed < 1 year ago | 30875 (1.2) | 253 (0.82) | 151 (0.49) | 6 (0.02) | 178 (0.58) | 3090 (10.01) |
| Diagnosed 1-4.9 years ago | 91267 (3.6) | 515 (0.56) | 285 (0.31) | 19 (0.02) | 307 (0.34) | 3495 (3.83) |
| Diagnosed ≥5 years ago | 243542 (9.5) | 1418 (0.58) | 781 (0.32) | 57 (0.02) | 771 (0.32) | 5598 (2.30) |
| **Reduced kidney function** |  |  |  |  |  |  |
| Estimated GFR 30-60 | 535850 (20.9) | 3695 (0.69) | 2268 (0.42) | 146 (0.03) | 2493 (0.47) | 13378 (2.50) |
| Estimated GFR <30 | 39509 (1.5) | 672 (1.70) | 498 (1.26) | 15 (0.04) | 589 (1.49) | 3271 (8.28) |
| **End-stage renal disease*** | 6279 (0.2) | 181 (2.88) | 141 (2.25) | 11 (0.18) | 117 (1.86) | 441 (7.02) |
| **Chronic Liver disease** | 24604 (1.0) | 236 (0.96) | 131 (0.53) | 7 (0.03) | 134 (0.54) | 926 (3.76) |
| **Stroke/dementia** | 187946 (7.3) | 2068 (1.10) | 1166 (0.62) | 54 (0.03) | 1331 (0.71) | 7462 (3.97) |
| **Other neurological disease** | 50577 (2.0) | 569 (1.13) | 309 (0.61) | 12 (0.02) | 380 (0.75) | 1850 (3.66) |
| **Solid organ transplant**** | 825 (0.0) | 14 (1.70) | 7 (0.85) | REDACTED | 7 (0.85) | 27 (3.27) |
| **Asplenia** | 6355 (0.2) | 46 (0.72) | 34 (0.54) | REDACTED | 17 (0.27) | 164 (2.58) |
| **Rheumatoid/Lupus/ Psoriasis** | 199014 (7.8) | 1225 (0.62) | 669 (0.34) | 65 (0.03) | 608 (0.31) | 3438 (1.73) |
| **Other immunosuppressive condition** | 5235 (0.2) | 52 (0.99) | 28 (0.53) | REDACTED | 28 (0.53) | 189 (3.61) |
| **Shielding***** | 903291 (35.2) | 6151 (0.68) | 3383 (0.37) | 281 (0.03) | 3255 (0.36) | 23722 (2.63) |

*End-stage renal disease includes on dialysis or having had a kidney transplant

**All solid organ transplants, excluding kidney

***Shielding includes organ transplant recipients, renal replacement therapy, haematological cancers, non-haematological cancers, immunodeficiencies/asplenia and severe respiratory conditions.

**Table A4. Hazard Ratios (HRs) for outcomes (a) evidence of SARS-CoV-2 infection recorded in primary care, (b) COVID-19 hospital admission, (c) COVID-19 ICU admission, (d) COVID-19 death and (e) non-COVID-19 death), stratified by age.**

(a) **Recorded SARS-CoV-2 infection**

|  | | | | **Recorded SARS-CoV-2 infection**  **HR (95% CI)** | | |
| --- | --- | --- | --- | --- | --- | --- |
|  | **N** | **Person years follow-up** | **Rate (per 100,000 person years)** | **Age-sex adjusted** | **+ Demographic adjusted*** | **+ Comorbidity adjusted**** |
| **Adults 65 years and under** | | | | | | |
| **Children in the household** |  |  |  |  |  |  |
| None | 17527 | 2862491 | 612.3 | 1.00 (ref) | 1.00 (ref) | 1.00 (ref) |
| Children aged 0-11 years | 8980 | 1285621 | 698.5 | 1.18 (1.14-1.21) | 1.02 (0.99-1.05) | 1.03 (1.00-1.06) |
| Children aged $\geq$12 years | 3356 | 426019 | 787.76 | 1.21 (1.16-1.26) | 1.08 (1.03-1.13) | 1.08 (1.04-1.13) |
| **Number of children aged 0-11 years in household** | | | | | | |
| None | 17527 | 2862491 | 612.3 | 1.00 (ref) | 1.00 (ref) | 1.00 (ref) |
| 1 child 0-11 years | 4919 | 698440 | 704.28 | 1.18 (1.13-1.22) | 1.04 (1.00-1.08) | 1.05 (1.01-1.09) |
| 2 children 0-11 years | 3058 | 451176 | 677.78 | 1.15 (1.10-1.21) | 1.01 (0.97-1.06) | 1.02 (0.98-1.07) |
| ⋝3 children 0-11 years | 1003 | 136006 | 737.47 | 1.25 (1.16-1.34) | 0.95 (0.88-1.03) | 0.97 (0.90-1.04) |
| **Adults over 65 years** | | | | | | |
| **Children in the household** |  |  |  |  |  |  |
| None | 11331 | 1233014 | 918.97 | 1.00 (ref) | 1.00 (ref) | 1.00 (ref) |
| Children aged 0-11 years | 354 | 28808 | 1228.82 | 1.41 (1.26-1.57) | 1.08 (0.96-1.22) | 1.07 (0.95-1.20) |
| Children aged $\geq$12 years | 141 | 14041 | 1004.21 | 1.11 (0.93-1.31) | 0.91 (0.76-1.08) | 0.91 (0.76-1.08) |
| **Number of children aged 0-11 years in household** | | | | | | |
| None | 11331 | 1233014 | 918.97 | 1.00 (ref) | 1.00 (ref) | 1.00 (ref) |
| 1 child 0-11 years | 206 | 17855 | 1153.72 | 1.33 (1.15-1.54) | 1.05 (0.91-1.22) | 1.04 (0.90-1.21) |
| 2 children 0-11 years | 108 | 8562 | 1261.38 | 1.45 (1.19-1.77) | 1.10 (0.89-1.35) | 1.09 (0.89-1.34) |
| ⋝3 children 0-11 years | 40 | 2391 | 1673.09 | 1.80 (1.31-2.48) | 1.21 (0.88-1.67) | 1.17 (0.84-1.62) |

*Demographic adjusted model: Adjusted for age, sex, ethnicity, number adults in household, IMD, BMI, smoking.

**Comorbidity-adjusted model: Adjusted for age, sex, ethnicity, number adults in household, IMD, BMI, smoking, hypertension or high blood pressure, chronic respiratory disease, asthma, cancer, chronic liver disease, stroke or dementia, other neurological disease, reduced kidney function, end-stage renal disease, solid organ transplant, asplenia, rheumatoid, lupus or psoriasis, other immunosuppressive condition.
**(b) COVID-19 hospital admissions**

|  | | | | **COVID-19 hospital admissions**  **HR (95% CI)** | | |
| --- | --- | --- | --- | --- | --- | --- |
|  | **N** | **Person years follow-up** | **Rate (per 100,000 person years)** | **Age-sex adjusted** | **+ Demographic adjusted*** | **+ Comorbidity adjusted**** |
| **Adults 65 years and under** | | | | | | |
| **Children in the household** |  |  |  |  |  |  |
| None | 3261 | 1406950 | 231.78 | 1.00 (ref) | 1.00 (ref) | 1.00 (ref) |
| Children aged 0-11 years | 1006 | 631072 | 159.41 | 1.12 (1.04-1.21) | 0.96 (0.88-1.03) | 0.97 (0.90-1.05) |
| Children aged $\geq$12 years | 509 | 209029 | 243.51 | 1.19 (1.08-1.31) | 1.06 (0.96-1.18) | 1.08 (0.98-1.19) |
| **Number of children aged 0-11 years in household** | | | | | | |
| None | 3261 | 1406950 | 231.78 | 1.00 (ref) | 1.00 (ref) | 1.00 (ref) |
| 1 child 0-11 years | 564 | 342883 | 164.49 | 1.10 (1.00-1.21) | 0.96 (0.87-1.05) | 0.97 (0.88-1.07) |
| 2 children 0-11 years | 319 | 221405 | 144.08 | 1.07 (0.95-1.21) | 0.93 (0.82-1.05) | 0.94 (0.83-1.06) |
| ⋝3 children 0-11 years | 123 | 66784 | 184.18 | 1.43 (1.19-1.72) | 1.04 (0.86-1.25) | 1.07 (0.89-1.29) |
| **Adults over 65 years** | | | | | | |
| **Children in the household** |  |  |  |  |  |  |
| None | 6224 | 607518 | 1024.5 | 1.00 (ref) | 1.00 (ref) | 1.00 (ref) |
| Children aged 0-11 years | 181 | 14216 | 1273.23 | 1.26 (1.08-1.46) | 0.99 (0.84-1.16) | 0.96 (0.82-1.13) |
| Children aged $\geq$12 years | 91 | 6922 | 1314.69 | 1.24 (1.01-1.52) | 1.03 (0.83-1.28) | 1.03 (0.83-1.27) |
| **Number of children aged 0-11 years in household** | | | | | | |
| None | 6224 | 607518 | 1024.5 | 1.00 (ref) | 1.00 (ref) | 1.00 (ref) |
| 1 child 0-11 years | 100 | 8811 | 1134.97 | 1.13 (0.93-1.38) | 0.91 (0.74-1.12) | 0.90 (0.73-1.10) |
| 2 children 0-11 years | 53 | 4224 | 1254.59 | 1.25 (0.95-1.65) | 0.97 (0.73-1.29) | 0.94 (0.71-1.25) |
| ⋝3 children 0-11 years | 28 | 1181 | 2371.81 | 2.15 (1.48-3.11) | 1.48 (1.02-2.16) | 1.41 (0.97-2.07) |

*Demographic adjusted model: Adjusted for age, sex, ethnicity, number adults in household, IMD, BMI, smoking.

**Comorbidity-adjusted model: Adjusted for age, sex, ethnicity, number adults in household, IMD, BMI, smoking, hypertension or high blood pressure, chronic respiratory disease, asthma, cancer, chronic liver disease, stroke or dementia, other neurological disease, reduced kidney function, end-stage renal disease, solid organ transplant, asplenia, rheumatoid, lupus or psoriasis, other immunosuppressive condition.

**c) COVID-19 ICU admission**

|  | | | | **COVID-19 ICU admission HR (95% CI)** | | |
| --- | --- | --- | --- | --- | --- | --- |
|  | **N** | **Person years follow-up** | **Rate (per 100,000 person years)** | **Age-sex adjusted** | **+Demographic adjusted*** | **+Comorbidity adjusted**** |
| **Adults 65 years and under** | | | | | | |
| **Children in the household** |  |  |  |  |  |  |
| None | 1037 | 2866026 | 36.18 | 1.00 (ref) | 1.00 (ref) | 1.00 (ref) |
| Children aged 0-11 years | 280 | 1287327 | 21.75 | 1.21 (1.04-1.41) | 0.96 (0.82-1.12) | 0.97 (0.83-1.13) |
| Children aged $\geq$12 years | 154 | 426669 | 36.09 | 1.22 (1.02-1.45) | 1.04 (0.87-1.25) | 1.06 (0.88-1.26) |
| **Number of children aged 0-11 years in household** | | | | | | |
| None | 1037 | 2866026 | 36.18 | 1.00 (ref) | 1.00 (ref) | 1.00 (ref) |
| 1 child 0-11 years | 169 | 699378 | 24.16 | 1.25 (1.05-1.49) | 1.01 (0.85-1.21) | 1.02 (0.86-1.22) |
| 2 children 0-11 years | 81 | 451766 | 17.93 | 1.08 (0.85-1.37) | 0.87 (0.68-1.10) | 0.88 (0.69-1.12) |
| ⋝3 children 0-11 years | 30 | 136183 | 22.03 | 1.42 (0.98-2.06) | 0.92 (0.63-1.34) | 0.94 (0.65-1.37) |
| **Adults over 65 years** | | | | | | |
| **Children in the household** |  |  |  |  |  |  |
| None | 634 | 1235141 | 51.33 | 1.00 (ref) | 1.00 (ref) | 1.00 (ref) |
| Children aged 0-11 years | 27 | 28872 | 93.52 | 1.34 (0.91-1.98) | 0.94 (0.62-1.43) | 0.94 (0.62-1.42) |
| Children aged $\geq$12 years | 14 | 14064 | 99.54 | 1.56 (0.92-2.65) | 1.20 (0.69-2.09) | 1.20 (0.69-2.09) |
| **Number of children aged 0-11 years in household** | | | | | | |
| None | 634 | 1235141 | 51.33 | 1.00 (ref) | 1.00 (ref) | 1.00 (ref) |
| 1 child 0-11 years | 15 | 17895 | 83.82 | 1.22 (0.73-2.04) | 0.89 (0.52-1.52) | 0.89 (0.52-1.51) |
| 2 children 0-11 years | 10 | 8580 | 116.56 | 1.64 (0.87-3.07) | 1.12 (0.59-2.14) | 1.12 (0.59-2.13) |
| ⋝3 children 0-11 years | REDACTED | 2397 | 83.44 | 1.18 (0.29-4.70) | 0.69 (0.17-2.84) | 0.69 (0.17-2.84) |

*Demographic adjusted model: Adjusted for age, sex, ethnicity, number adults in household, IMD, BMI, smoking.

### **Comorbidity-adjusted model: Adjusted for age, sex, ethnicity, number adults in household, IMD, BMI, smoking, hypertension or high blood pressure, chronic respiratory disease, asthma, cancer, chronic liver disease, stroke or dementia, other neurological disease, reduced kidney function, end-stage renal disease, solid organ transplant, asplenia, rheumatoid, lupus or psoriasis, other immunosuppressive condition.

### **d) COVID-19 death**

|  | | | | **COVID-19 death HR (95% CI)** | | |
| --- | --- | --- | --- | --- | --- | --- |
|  | **N** | **Person years follow-up** | **Rate**  **(per 100,000 person years)** | **Age-sex adjusted** | **+Demographic adjusted*** | **+Comorbidity adjusted**** |
| **Adults 65 years and under** | | | | | | |
| **Children in the household** |  |  |  |  |  |  |
| None | 953 | 2866242 | 33.25 | 1.00 (ref) | 1.00 (ref) | 1.00 (ref) |
| Children aged 0-11 years | 136 | 1287389 | 10.56 | 0.87 (0.71-1.06) | 0.74 (0.60-0.90) | 0.75 (0.62-0.92) |
| Children aged $\geq$12 years | 84 | 426705 | 19.69 | 0.89 (0.70-1.12) | 0.83 (0.66-1.05) | 0.85 (0.67-1.07) |
| **Number of children aged 0-11 years in household** | | | | | | |
| None | 953 | 2866242 | 33.25 | 1.00 (ref) | 1.00 (ref) | 1.00 (ref) |
| 1 child 0-11 years | 90 | 699414 | 12.87 | 0.95 (0.76-1.20) | 0.82 (0.66-1.03) | 0.84 (0.67-1.05) |
| 2 children 0-11 years | 34 | 451785 | 7.53 | 0.70 (0.48-1.00) | 0.60 (0.42-0.86) | 0.61 (0.43-0.88) |
| ⋝3 children 0-11 years | 12 | 136190 | 8.81 | 0.86 (0.48-1.55) | 0.62 (0.35-1.12) | 0.64 (0.36-1.16) |
| **Adults over 65 years** | | | | | | |
| **Children in the household** |  |  |  |  |  |  |
| None | 6148 | 1235225 | 497.72 | 1.00 (ref) | 1.00 (ref) | 1.00 (ref) |
| Children aged 0-11 years | 138 | 28874 | 477.93 | 1.19 (1.00-1.41) | 0.86 (0.72-1.03) | 0.85 (0.71-1.01) |
| Children aged $\geq$12 years | 66 | 14066 | 469.21 | 1.04 (0.82-1.34) | 0.82 (0.64-1.06) | 0.82 (0.64-1.05) |
| **Number of children aged 0-11 years in household** | | | | | | |
| None | 6148 | 1235225 | 497.72 | 1.00 (ref) | 1.00 (ref) | 1.00 (ref) |
| 1 child 0-11 years | 82 | 17897 | 458.18 | 1.15 (0.92-1.43) | 0.86 (0.68-1.08) | 0.85 (0.67-1.06) |
| 2 children 0-11 years | 38 | 8580 | 442.87 | 1.12 (0.81-1.54) | 0.80 (0.58-1.11) | 0.79 (0.57-1.09) |
| ⋝3 children 0-11 years | 18 | 2397 | 750.9 | 1.68 (1.06-2.67) | 1.06 (0.66-1.69) | 1.00 (0.62-1.60) |

*Demographic adjusted model: Adjusted for age, sex, ethnicity, number adults in household, IMD, BMI, smoking.

**Comorbidity-adjusted model: Adjusted for age, sex, ethnicity, number adults in household, IMD, BMI, smoking, hypertension or high blood pressure, chronic respiratory disease, asthma, cancer, chronic liver disease, stroke or dementia, other neurological disease, reduced kidney function, end-stage renal disease, solid organ transplant, asplenia, rheumatoid, lupus or psoriasis, other immunosuppressive condition.
**e) non-COVID-19 deaths**

|  | | | | **Non-COVID-19 Death HR (95% CI)** | | |
| --- | --- | --- | --- | --- | --- | --- |
|  | **N** | **Person years follow-up** | **Rate**  **(per 100,000 person years)** | **Age-sex adjusted** | **+Demographic adjusted*** | **+Comorbidity adjusted**** |
| **Adults 65 years and under** | | | | | | |
| **Children in the household** |  |  |  |  |  |  |
| None | 6660 | 2866242 | 232.36 | 1.00 (ref) | 1.00 (ref) | 1.00 (ref) |
| Children aged 0-11 years | 697 | 1287389 | 54.14 | 0.55 (0.51-0.60) | 0.65 (0.60-0.71) | 0.68 (0.62-0.74) |
| Children aged $\geq$12 years | 431 | 426705 | 101.01 | 0.60 (0.54-0.66) | 0.72 (0.65-0.79) | 0.73 (0.66-0.81) |
| **Number of children aged 0-11 years in household** | | | | | | |
| None | 6660 | 2866242 | 232.36 | 1.00 (ref) | 1.00 (ref) | 1.00 (ref) |
| 1 child 0-11 years | 425 | 699414 | 60.77 | 0.57 (0.52-0.64) | 0.66 (0.60-0.74) | 0.69 (0.62-0.76) |
| 2 children 0-11 years | 215 | 451785 | 47.59 | 0.53 (0.46-0.61) | 0.65 (0.56-0.75) | 0.67 (0.58-0.77) |
| ⋝3 children 0-11 years | 57 | 136190 | 41.85 | 0.51 (0.39-0.66) | 0.58 (0.44-0.76) | 0.63 (0.48-0.82) |
| **Adults over 65 years** | | | | | | |
| **Children in the household** |  |  |  |  |  |  |
| None | 36531 | 1235225 | 2957.44 | 1.00 (ref) | 1.00 (ref) | 1.00 (ref) |
| Children aged 0-11 years | 614 | 28874 | 2126.44 | 1.04 (0.96-1.12) | 0.98 (0.90-1.06) | 0.98 (0.90-1.07) |
| Children aged$\geq$12 years | 334 | 14066 | 2374.5 | 1.00 (0.90-1.12) | 0.96 (0.86-1.07) | 0.96 (0.86-1.08) |
| **Number of children aged 0-11 years in household** | | | | | | |
| None | 36531 | 1235225 | 2957.44 | 1.00 (ref) | 1.00 (ref) | 1.00 (ref) |
| 1 child 0-11 years | 383 | 17897 | 2140.03 | 1.04 (0.94-1.15) | 0.98 (0.88-1.09) | 0.98 (0.88-1.09) |
| 2 children 0-11 years | 173 | 8580 | 2016.22 | 1.00 (0.86-1.17) | 0.95 (0.82-1.11) | 0.98 (0.84-1.14) |
| ⋝3 children 0-11 years | 58 | 2397 | 2419.56 | 1.12 (0.86-1.45) | 1.03 (0.79-1.34) | 0.98 (0.75-1.27) |

*Demographic adjusted model: Adjusted for age, sex, ethnicity, number adults in household, IMD, BMI, smoking.

**Comorbidity-adjusted model: Adjusted for age, sex, ethnicity, number adults in household, IMD, BMI, smoking, hypertension or high blood pressure, chronic respiratory disease, asthma, cancer, chronic liver disease, stroke or dementia, other neurological disease, reduced kidney function, end-stage renal disease, solid organ transplant, asplenia, rheumatoid, lupus or psoriasis, other immunosuppressive condition.

**Table A5. Hazard Ratios (HRs) for threadworm between 2018 to 2019, stratified by age**

|  | | | | **Worms HR (95% CI)** | | |
| --- | --- | --- | --- | --- | --- | --- |
|  | **N**  **(column %)** | **Person years follow-up** | **Rate**  **per 100,000 person years)** | **Age-sex adjusted** | **+ Demographic adjusted*** | **+ Comorbidity adjusted**** |
| **Adults 65 years and under** | | | | | | |
| **Children in the household** |  |  |  |  |  |  |
| None | 462 | 5587465 | 8.27 | 1.00 (ref) | 1.00 (ref) | 1.00 (ref) |
| Children aged 0-11 years | 620 | 2395112 | 25.89 | 2.56 (2.23-2.94) | 2.52 (2.19-2.91) | 2.53 (2.19-2.92) |
| Children aged $\geq$12 years | 81 | 766691 | 10.56 | 1.28 (1.00-1.64) | 1.31 (1.02-1.68) | 1.32 (1.03-1.70) |
| **Number of children aged 1-<11 years in household** | | | | | | |
| None | 462 | 5587465 | 8.27 | 1.00 (ref) | 1.00 (ref) | 1.00 (ref) |
| 1 child 0-11 years | 238 | 1288807 | 18.47 | 1.88 (1.59-2.24) | 1.87 (1.57-2.23) | 1.87 (1.57-2.23) |
| 2 children 0-11 years | 258 | 851008 | 30.32 | 3.05 (2.55-3.65) | 3.06 (2.55-3.67) | 3.07 (2.56-3.69) |
| ⋝3 children 0-11 years | 124 | 255297 | 48.57 | 4.67 (3.73-5.84) | 4.54 (3.61-5.71) | 4.55 (3.61-5.72) |
| **Adults over 65 years** | | | | | | |
| **Children in the household** |  |  |  |  |  |  |
| None | 75 | 2155971 | 3.48 | 1.00 (ref) | 1.00 (ref) | 1.00 (ref) |
| Children aged 0-11 years | 6 | 44597 | 13.45 | 3.47 (1.28-9.42) | 3.25 (1.05-10.05) | 2.34 (0.55-9.97) |
| Children aged $\geq$12 years | REDACTED | 22686 | 8.82 | 2.39 (0.60-9.58) | 2.34 (0.52-10.48) | 1.24 (0.15-10.54) |
| **Number of children aged 1-<11 years in household** |  |  |  |  |  |  |
| None | 75 | 2155971 | 3.48 | 1.00 (ref) | 1.00 (ref) | 1.00 (ref) |
| 1 child 0-11 years | REDACTED | 27539 | 7.26 | 1.89 (0.25-14.27) | 1.88 (0.24-14.64) | 1.98 (0.25-15.83) |
| 2 children 0-11 years | REDACTED | 13373 | 14.96 | 3.86 (0.93-15.91) | 3.69 (0.82-16.73) | 1.95 (0.22-16.94) |
| ⋝3 children 0-11 years | REDACTED | 3685 | 54.27 | 13.61 (3.26-56.82) | 12.10 (2.17-67.45) | 6.25 (0.66-59.52) |

*Demographic adjusted model: Adjusted for age, sex, ethnicity, number adults in household, IMD, BMI, smoking.

**Comorbidity-adjusted model: Adjusted for age, sex, ethnicity, number adults in household, IMD, BMI, smoking, hypertension or high blood pressure, chronic respiratory disease, asthma, cancer, chronic liver disease, stroke or dementia, other neurological disease, reduced kidney function, end-stage renal disease, solid organ transplant, asplenia, rheumatoid, lupus or psoriasis, other immunosuppressive condition.

### **Table A6: Hazard Ratios (HRs) and 95% confidence intervals (CIs) for COVID-19 outcomes among those 65 years and under, compare results of complete case analysis and using multiple imputation to account for ethnicity.**

|  | **Evidence of SARS-CoV-2 infection in primary care** | | **Hospital admission** | | **ICU admission** | | **Death from COVID-19** | |
| --- | --- | --- | --- | --- | --- | --- | --- | --- |
|  | Complete case* | Multiple imputation | Complete case* | Multiple imputation | Complete case* | Multiple imputation | Complete case* | Multiple imputation |
| None | 1.00 (ref) | 1.00 (ref) | 1.00 (ref) | 1.00 (ref) | 1.00 (ref) | 1.00 (ref) | 1.00 (ref) | 1.00 (ref) |
| Children aged 0-11 years | 1.03 (1.00-1.06) | 1.07 (1.04-1.10) | 0.97 (0.90-1.05) | 1.00 (0.93-1.07) | 0.97 (0.83-1.13) | 1.04 (0.90-1.19) | 0.75 (0.62-0.92) | 0.79 (0.66-0.94) |
| Children aged $\geq$12 years | 1.08 (1.04-1.13) | 1.10 (1.06-1.14) | 1.08 (0.98-1.19) | 1.09 (0.99-1.19) | 1.06 (0.88-1.26) | 1.13 (0.96-1.32) | 0.85 (0.67-1.07) | 0.91 (0.74-1.12) |

*Comorbidity-adjusted model: Adjusted for age, sex, ethnicity, number adults in household, IMD, BMI, smoking, hypertension or high blood pressure, chronic respiratory disease, asthma, cancer, chronic liver disease, stroke or dementia, other neurological disease, reduced kidney function, end-stage renal disease, solid organ transplant, asplenia, rheumatoid, lupus or psoriasis, other immunosuppressive condition.

**Table A7: Bias-adjusted hazard ratios accounting for high-risk occupation for the association between living in a household with children aged 0-11 years and recorded SARS-CoV-2 infection among adults aged ≤ 65 years: (Main analysis comorbidity adjusted HR 1.03, 95% CI 1.00-1.06)**

| Assumed risk ratio between high-risk work and infection with SARS-CoV-2 | Assumed prevalence of high-risk workers among unexposed | Assumed prevalence of high-risk workers among exposed | | |
| --- | --- | --- | --- | --- |
|  |  | 0.25 | 0.3 | 0.35 |
| 1.3 | 0.25 | 1.03 (1.00-1.06) | 1.02 (0.99-1.05) | 1.00 (0.97-1.03) |
|  | 0.3 | 1.04 (1.01-1.07) | 1.03 (1.00-1.06) | 1.02 (0.99-1.05) |
|  | 0.35 | 1.06 (1.03-1.09) | 1.04 (1.01-1.07) | 1.03 (1.00-1.06) |
| 1.6 | 0.25 | 1.03 (1.00-1.06) | 1.00 (0.97-1.03) | 0.98 (0.95-1.01) |
|  | 0.3 | 1.06 (1.03-1.09) | 1.03 (1.00-1.06) | 1.00 (0.98-1.03) |
|  | 0.35 | 1.08 (1.05-1.12) | 1.06 (1.03-1.09) | 1.03 (1.00-1.06) |
| 2 | 0.25 | 1.03 (1.00-1.06) | 0.99 (0.96-1.02) | 0.95 (0.93-0.98) |
|  | 0.3 | 1.07 (1.04-1.10) | 1.03 (1.00-1.06) | 0.99 (0.96-1.02) |
|  | 0.35 | 1.11 (1.08-1.14) | 1.07 (1.04-1.10) | 1.03 (1.00-1.06) |

**Table A8: Bias-adjusted hazard ratios accounting for high-risk occupation for the association between living in a household with children aged ≥12 years and recorded SARS-CoV-2 infection among adults aged ≤ 65 years: (Main analysis comorbidity adjusted HR 1.08, 95% CI 1.04-1.13)**

| Assumed risk ratio between high-risk work and infection with SARS-CoV-2 | Assumed prevalence of high-risk workers among unexposed | Assumed prevalence of high-risk workers among exposed | | |
| --- | --- | --- | --- | --- |
|  |  | 0.25 | 0.3 | 0.35 |
| 1.3 | 0.25 | 1.08 (1.04-1.13) | 1.07 (1.03-1.11) | 1.05 (1.01-1.10) |
|  | 0.3 | 1.10 (1.05-1.15) | 1.08 (1.04-1.13) | 1.07 (1.03-1.11) |
|  | 0.35 | 1.11 (1.07-1.16) | 1.09 (1.05-1.15) | 1.08 (1.04-1.13) |
| 1.6 | 0.25 | 1.08 (1.04-1.13) | 1.05 (1.01-1.10) | 1.03 (0.99-1.07) |
|  | 0.3 | 1.11 (1.07-1.16) | 1.08 (1.04-1.13) | 1.05 (1.01-1.10) |
|  | 0.35 | 1.14 (1.09-1.19) | 1.11 (1.07-1.16) | 1.08 (1.04-1.13) |
| 2 | 0.25 | 1.08 (1.04-1.13) | 1.04 (1.00-1.09) | 1.00 (0.96-1.05) |
|  | 0.3 | 1.12 (1.08-1.18) | 1.08 (1.04-1.13) | 1.04 (1.00-1.09) |
|  | 0.35 | 1.17 (1.12-1.22) | 1.12 (1.08-1.17) | 1.08 (1.04-1.13) |

**Table A9: Bias-adjusted hazard ratios accounting for high-risk occupation for the association between living in a household with children aged 0-11 years and Covid-19 hospital admission among adults aged ≤ 65 years: (Main analysis comorbidity adjusted HR 0.97, 95% CI 0.90-1.05)**

| Assumed risk ratio between high-risk work and infection with SARS-CoV-2 | Assumed prevalence of high-risk workers among unexposed | Assumed prevalence of high-risk workers among exposed | | |
| --- | --- | --- | --- | --- |
|  |  | 0.25 | 0.3 | 0.35 |
| 1.3 | 0.25 | 0.97 (0.90-1.05) | 0.96 (0.89-1.04) | 0.94 (0.88-1.02) |
|  | 0.3 | 0.98 (0.91-1.06) | 0.97 (0.90-1.05) | 0.96 (0.89-1.04) |
|  | 0.35 | 1.00 (0.93-1.08) | 0.98 (0.91-1.06) | 0.97 (0.90-1.05) |
| 1.6 | 0.25 | 0.97 (0.90-1.05) | 0.95 (0.88-1.02) | 0.92 (0.86-1.00) |
|  | 0.3 | 1.00 (0.92-1.08) | 0.97 (0.90-1.05) | 0.95 (0.88-1.02) |
|  | 0.35 | 1.02 (0.95-1.10) | 0.99 (0.92-1.08) | 0.97 (0.90-1.05) |
| 2 | 0.25 | 0.97 (0.90-1.05) | 0.93 (0.87-1.01) | 0.90 (0.83-0.97) |
|  | 0.3 | 1.01 (0.94-1.09) | 0.97 (0.90-1.05) | 0.93 (0.87-1.01) |
|  | 0.35 | 1.05 (0.97-1.13) | 1.01 (0.93-1.09) | 0.97 (0.90-1.05) |

**Table A10: Bias-adjusted hazard ratios accounting for high-risk occupation for the association between living in a household with children aged ≥12 years and Covid-19 hospital admission among adults aged ≤ 65 years (Main analysis comorbidity adjusted HR 1.08, 95% CI 0.98-1.19)**

| Assumed risk ratio between high-risk work and infection with SARS-CoV-2 | Assumed prevalence of high-risk workers among unexposed | Assumed prevalence of high-risk workers among exposed | | |
| --- | --- | --- | --- | --- |
|  |  | 0.25 | 0.3 | 0.35 |
| 1.3 | 0.25 | 1.06 (0.88-1.26) | 1.05 (0.87-1.24) | 1.03 (0.86-1.23) |
|  | 0.3 | 1.07 (0.89-1.28) | 1.06 (0.88-1.26) | 1.05 (0.87-1.24) |
|  | 0.35 | 1.09 (0.90-1.30) | 1.07 (0.89-1.28) | 1.06 (0.88-1.26) |
| 1.6 | 0.25 | 1.06 (0.88-1.26) | 1.03 (0.86-1.23) | 1.01 (0.84-1.20) |
|  | 0.3 | 1.09 (0.90-1.29) | 1.06 (0.88-1.26) | 1.03 (0.86-1.23) |
|  | 0.35 | 1.12 (0.93-1.33) | 1.09 (0.90-1.29) | 1.06 (0.88-1.26) |
| 2 | 0.25 | 1.06 (0.88-1.26) | 1.02 (0.85-1.21) | 0.98 (0.81-1.17) |
|  | 0.3 | 1.10 (0.92-1.31) | 1.06 (0.88-1.26) | 1.02 (0.85-1.21) |
|  | 0.35 | 1.14 (0.95-1.36) | 1.10 (0.91-1.31) | 1.06 (0.88-1.26) |

**Table A11: Bias-adjusted hazard ratios accounting for high-risk occupation for the association between living in a household with children aged 0-11 years and Covid-19 ICU admission among adults aged ≤ 65 years: (Main analysis comorbidity adjusted HR 0.97, 95% CI 0.83-1.13)**

| Assumed risk ratio between high-risk work and infection with SARS-CoV-2 | Assumed prevalence of high-risk workers among unexposed | Assumed prevalence of high-risk workers among exposed | | |
| --- | --- | --- | --- | --- |
|  |  | 0.25 | 0.3 | 0.35 |
| 1.3 | 0.25 | 0.97 (0.83-1.13) | 0.96 (0.82-1.11) | 0.94 (0.81-1.10) |
|  | 0.3 | 0.98 (0.84-1.15) | 0.97 (0.83-1.13) | 0.96 (0.82-1.11) |
|  | 0.35 | 1.00 (0.85-1.16) | 0.98 (0.84-1.15) | 0.97 (0.83-1.13) |
| 1.6 | 0.25 | 0.97 (0.83-1.13) | 0.95 (0.81-1.10) | 0.92 (0.79-1.07) |
|  | 0.3 | 1.00 (0.85-1.16) | 0.97 (0.83-1.13) | 0.95 (0.81-1.10) |
|  | 0.35 | 1.02 (0.87-1.19) | 0.99 (0.85-1.16) | 0.97 (0.83-1.13) |
| 2 | 0.25 | 0.97 (0.83-1.13) | 0.93 (0.80-1.09) | 0.90 (0.77-1.05) |
|  | 0.3 | 1.01 (0.86-1.18) | 0.97 (0.83-1.13) | 0.93 (0.80-1.09) |
|  | 0.35 | 1.05 (0.90-1.22) | 1.01 (0.86-1.17) | 0.97 (0.83-1.13) |

**Table A12: Bias-adjusted hazard ratios accounting for high-risk occupation for the association between living in a household with children aged ≥12 years and Covid-19 ICU admission among adults aged ≤ 65 years: (Main analysis comorbidity adjusted HR 1.06, 95% CI 0.88-1.26)**

| Assumed risk ratio between high-risk work and infection with SARS-CoV-2 | Assumed prevalence of high-risk workers among unexposed | Assumed prevalence of high-risk workers among exposed | | |
| --- | --- | --- | --- | --- |
|  |  | 0.25 | 0.3 | 0.35 |
| 1.3 | 0.25 | 1.06 (0.88-1.26) | 1.05 (0.87-1.24) | 1.03 (0.86-1.23) |
|  | 0.3 | 1.07 (0.89-1.28) | 1.06 (0.88-1.26) | 1.05 (0.87-1.24) |
|  | 0.35 | 1.09 (0.90-1.30) | 1.07 (0.89-1.28) | 1.06 (0.88-1.26) |
| 1.6 | 0.25 | 1.06 (0.88-1.26) | 1.03 (0.86-1.23) | 1.01 (0.84-1.20) |
|  | 0.3 | 1.09 (0.90-1.29) | 1.06 (0.88-1.26) | 1.03 (0.86-1.23) |
|  | 0.35 | 1.12 (0.93-1.33) | 1.09 (0.90-1.29) | 1.06 (0.88-1.26) |
| 2 | 0.25 | 1.06 (0.88-1.26) | 1.02 (0.85-1.21) | 0.98 (0.81-1.17) |
|  | 0.3 | 1.10 (0.92-1.31) | 1.06 (0.88-1.26) | 1.02 (0.85-1.21) |
|  | 0.35 | 1.14 (0.95-1.36) | 1.10 (0.91-1.31) | 1.06 (0.88-1.26) |

**Table A13: Bias-adjusted hazard ratios accounting for high-risk occupation for the association between living in a household with children aged 0-11 years and Covid-19 death among adults aged ≤ 65 years: (Main analysis comorbidity adjusted HR 0.75, 95% CI 0.62-0.92)**

| Assumed risk ratio between high-risk work and infection with SARS-CoV-2 | Assumed prevalence of high-risk workers among unexposed | Assumed prevalence of high-risk workers among exposed | | |
| --- | --- | --- | --- | --- |
|  |  | 0.25 | 0.3 | 0.35 |
| 1.3 | 0.25 | 0.75 (0.62-0.92) | 0.74 (0.61-0.91) | 0.73 (0.60-0.90) |
|  | 0.3 | 0.76 (0.63-0.93) | 0.75 (0.62-0.92) | 0.74 (0.61-0.91) |
|  | 0.35 | 0.77 (0.64-0.95) | 0.76 (0.63-0.93) | 0.75 (0.62-0.92) |
| 1.6 | 0.25 | 0.75 (0.62-0.92) | 0.73 (0.60-0.90) | 0.71 (0.59-0.87) |
|  | 0.3 | 0.77 (0.64-0.94) | 0.75 (0.62-0.92) | 0.73 (0.60-0.90) |
|  | 0.35 | 0.79 (0.65-0.97) | 0.77 (0.64-0.94) | 0.75 (0.62-0.92) |
| 2 | 0.25 | 0.75 (0.62-0.92) | 0.72 (0.60-0.88) | 0.69 (0.57-0.85) |
|  | 0.3 | 0.78 (0.64-0.96) | 0.75 (0.62-0.92) | 0.72 (0.60-0.89) |
|  | 0.35 | 0.81 (0.67-0.99) | 0.78 (0.64-0.96) | 0.75 (0.62-0.92) |

**Table A14: Bias-adjusted hazard ratios accounting for high-risk occupation for the association between living in a household with children aged ≥12 years and Covid-19 death among adults aged ≤ 65 years: (Main analysis comorbidity adjusted HR 0.85, 95% CI 0.67-1.07)**

| Assumed risk ratio between high-risk work and infection with SARS-CoV-2 | Assumed prevalence of high-risk workers among unexposed | Assumed prevalence of high-risk workers among exposed | | |
| --- | --- | --- | --- | --- |
|  |  | 0.25 | 0.3 | 0.35 |
| 1.3 | 0.25 | 1.08 (0.98-1.19) | 1.07 (0.97-1.17) | 1.05 (0.95-1.16) |
|  | 0.3 | 1.10 (0.99-1.21) | 1.08 (0.98-1.19) | 1.07 (0.97-1.17) |
|  | 0.35 | 1.11 (1.01-1.22) | 1.09 (0.99-1.21) | 1.08 (0.98-1.19) |
| 1.6 | 0.25 | 1.08 (0.98-1.19) | 1.05 (0.96-1.16) | 1.03 (0.93-1.13) |
|  | 0.3 | 1.11 (1.01-1.22) | 1.08 (0.98-1.19) | 1.05 (0.96-1.16) |
|  | 0.35 | 1.14 (1.03-1.25) | 1.11 (1.00-1.22) | 1.08 (0.98-1.19) |
| 2 | 0.25 | 1.08 (0.98-1.19) | 1.04 (0.94-1.14) | 1.00 (0.91-1.10) |
|  | 0.3 | 1.12 (1.02-1.24) | 1.08 (0.98-1.19) | 1.04 (0.94-1.15) |
|  | 0.35 | 1.17 (1.06-1.29) | 1.12 (1.02-1.24) | 1.08 (0.98-1.19) |

**Supplementary methods**

### **Algorithm to identify households in TPP, using addresses registered in patient record**

### 1) Get all address details for fully registered patients

### - Find all permanent patient addresses that were active on 01 Feb 2020 or recorded since

### - Combine the house name/number, remove punctuation (commas, full stops, apostrophes), double spaces and leading/trailing whitespace from house name/number and road

### - Replace street, lane etc with the abbreviation (street -> St, lane -> Ln, place -> Pl, avenue -> Ave, road -> Rd, close -> Cl, drive -> Dr)

### - Find registrations for patients that were active on or since 01 Feb 2020 (making sure the patient was alive on 01 Feb 2020)

### - Filter the patient addresses based on those registrations

### 2) Build an Address table using the distinct address fields

### - For each distinct (house name/number + road + post code), assign a unique Address_ID

### 3) Set the Address_ID on each individual patient address

### - Join to Address table from 2) on house name/number + road + post code

### 4) End addresses that started before the property was sold

### - Import land registry property sales data ([www.gov.uk/government/statistical-data-sets/price-paid-data-downloads](http://www.gov.uk/government/statistical-data-sets/price-paid-data-downloads))

### - Apply the same processing as 1) to the address fields

### - Set address IDs on the land registry house sales data using the same Address table from 2) joining on house name/number + road + post code

### - End all patient addresses at properties where the patient address start date was before the property was sold (under the assumption that in the majority of cases that means the occupants moved out)

### - If a patient address has an unset start date (imported data), use the registration start date for this step

### 5) Get the latest "active" patient address per patient

### - Filter to those which were active on 01 Feb 2020 or recorded since (need to do this again now we've ended some based on property sales)

### - Filter to the latest per patient

### 6) Create the HouseholdMember table

### - Insert a row for each patient address from 5), using the Address_ID as the Household_ID (one household per address)

### 7) Create the Household table

### - Select the distinct Address_IDs as the Household_ID into the Household table (one household per address)

### - Set the size based on the number of members

### - Use the PotentialCareHomeAddress table to set the CareHome flag

### - Check the address fields to identify NFA / Unknown addresses (e.g. postcode "ZZ99 ...", house name "NFA", road "Unknown", ...)

**Table of primary care codes used as evidence of SARS-CoV-2 infection in primary care with links to codelists**

| **Evidence** | **Link to OpenSAFELY codelists** |
| --- | --- |
| Clinical diagnosis of COVID-19 | https://codelists.opensafely.org/codelist/opensafely/covid-identification-in-primary-care-probable-covid-clinical-code/2020-07-16/ |
| Positive swab test for SARS-CoV-2 | https://codelists.opensafely.org/codelist/opensafely/covid-identification-in-primary-care-probable-covid-positive-test/2020-07-16/ |
| Sequelae of COVID-19 | https://codelists.opensafely.org/codelist/opensafely/covid-identification-in-primary-care-probable-covid-sequelae/2020-07-16/ |

**Table of covariate definitions**

| **Covariate** | **Link to OpenSAFELY codelists** |
| --- | --- |
| Obesity | Body mass index (BMI) was ascertained within the 10 years prior to 1 Feb 2020 and recorded when the patient was over 16 years old. Grouped using categories derived from the World Health Organisation classification of BMI: no evidence of obesity <30 kg/m^2^; obese I 30-34.9; obese II 35-39.9; obese III 40+ |
| Smoking | Grouped into current, former and never smokers. |
| Ethnicity | Categorised as a five-level variable, White, Mixed, South Asian, Black or Other |
| Age | age groups were: 18-29, 30-39, 40-49, 50-59, 60-69, 70-79, 80+ years. |
| Total number of adults in the household | Categorised as 1, 2, and 3 or more. |
| Region | We used the Sustainability and Transformation Partnership area (STP, an NHS administrative grouping) of the patient’s general practice as a marker of region. |
| Shielding patients | Identified at study start as having one or more of (ever); organ transplant, renal replacement therapy, haematological cancers, non-haematological cancers, immunodeficiencies/asplenia and chronic respiratory disease. |

**Table of chronic comorbidities with details and links to codelists**

| **Comorbidity** | **Details** | **Link to OpenSAFELY codelists** | **Time frame codes searched** |
| --- | --- | --- | --- |
| Asthma | Asthma code AND no codes indicating chronic obstructive pulmonary disease | https://codelists.opensafely.org/codelist/opensafely/asthma-diagnosis/2020-04-15/ | Ever diagnosed prior to study start date |
| Chronic Respiratory Disease | Including chronic obstructive pulmonary disease, fibrosing lung disease, bronchiectasis or cystic fibrosis | https://codelists.opensafely.org/codelist/opensafely/chronic-respiratory-disease/2020-04-10/ | Ever diagnosed prior to study start date |
| Chronic heart disease | Including chronic heart failure, ischaemic heart disease, and severe valve or congenital heart disease likely to require lifelong follow-up | https://codelists.opensafely.org/codelist/opensafely/chronic-cardiac-disease-snomed/2020-04-08-draft/ | Ever diagnosed prior to study start date |
| Diabetes mellitus | Defined using a combination of diabetes diagnosis codes, anti-diabetes drugs and HBA1C records were used to define diabetes status. | <https://codelists.opensafely.org/codelist/opensafely/diabetes-unknown-type/2020-06-29/>  <https://codelists.opensafely.org/codelist/opensafely/type-1-diabetes/2020-06-29/>  <https://codelists.opensafely.org/codelist/opensafely/type-2-diabetes/2020-06-29/>  <https://codelists.opensafely.org/codelist/opensafely/antidiabetic-drugs/2020-07-16/>  https://codelists.opensafely.org/codelist/opensafely/insulin-medication/2020-04-26/ | Ever diagnosed prior to study start date |
| Chronic liver disease | Including all chronic viral hepatitis disease, signs of cirrhosis such as oesophageal varices and liver transplant (recipient only) | https://codelists.opensafely.org/codelist/opensafely/chronic-liver-disease/2020-06-02/ | Ever diagnosed prior to study start date |
| Stroke/dementia | Diagnoses of stroke or dementia. | <https://codelists.opensafely.org/codelist/opensafely/stroke-updated/2020-06-02/>  https://codelists.opensafely.org/codelist/opensafely/dementia/2020-04-22/ | Ever diagnosed prior to study start date |
| Other chronic neurological diseases | Including conditions in which respiratory function may be compromised, such as motor neurone disease, myasthenia gravis, multiple sclerosis, Parkinson's disease, cerebral palsy, quadriplegia or hemiplegia and progressive cerebellar disease. | https://codelists.opensafely.org/codelist/opensafely/other-neurological-conditions/2020-06-02/ | Ever diagnosed prior to study start date |
| Common autoimmune diseases | Rheumatoid Arthritis, Systemic Lupus Erythematosus or psoriasis | https://codelists.opensafely.org/codelist/opensafely/ra-sle-psoriasis/2020-04-14/ | Ever diagnosed prior to study start date |
| Solid organ transplant | Codes indicating other organ transplant (kidney is grouped with renal replacement therapy below) | https://codelists.opensafely.org/codelist/opensafely/other-organ-transplant/2020-07-15/ | Ever diagnosed prior to study start date |
| Asplenia | Including splenectomy or a spleen dysfunction, including sickle cell disease. | https://codelists.opensafely.org/codelist/opensafely/asplenia/2020-06-02/ | Ever diagnosed prior to study start date |
| Immunosuppression (including permanent immunodeficiency such as HIV, sickle cell disease, other immunosuppressive conditions and temporary immunodeficiency) | Other immunosuppressive conditions included human immunodeficiency virus or a condition inducing permanent immunodeficiency ever diagnosed, or aplastic anaemia or temporary immunodeficiency recorded within the last year. | <https://codelists.opensafely.org/codelist/opensafely/hiv/2020-07-13/>  <https://codelists.opensafely.org/codelist/opensafely/permanent-immunosuppression/2020-06-02/>  <https://codelists.opensafely.org/codelist/opensafely/sickle-cell-disease/2020-04-14/>  <https://codelists.opensafely.org/codelist/opensafely/temporary-immunosuppression/2020-04-24/>  https://codelists.opensafely.org/codelist/opensafely/aplastic-anaemia/2020-04-24/ | Ever diagnosed prior to study start date |
| Cancer (haematological or non haematological) |  | <https://codelists.opensafely.org/codelist/opensafely/haematological-cancer/2020-04-15/>  <https://codelists.opensafely.org/codelist/opensafely/cancer-excluding-lung-and-haematological/2020-04-15/>  https://codelists.opensafely.org/codelist/opensafely/lung-cancer/2020-04-15/ | Cancer was grouped by time since the first diagnosis (within the last year; between 1 and 4.9 years ago; more than 5 years ago). |
| Estimated GFR (eGFR) | Kidney function was ascertained from the most recent serum creatinine measurement, and was converted into eGFR using the CKD-EPI equation, with reduced kidney function grouped into eGFR 30–59.9 or <30 ml/min/1.73m2. No serum creatinine measurement was categorized as eGFR>60mls.min/1.73m2 |  | Ever record prior to study start date |
| *End-stage renal disease includes | On dialysis or having had a kidney transplant ever. | <https://codelists.opensafely.org/codelist/opensafely/dialysis/2020-07-16/>  https://codelists.opensafely.org/codelist/opensafely/kidney-transplant/2020-07-15/ | Ever diagnosis prior to study start date |
| High blood pressure or diagnosis of hypertension | A previous coded diagnosis of hypertension or the most recent recording indicating systolic blood pressure ≥140 mmHg or diastolic blood pressure ≥90 mmHg | https://codelists.opensafely.org/codelist/opensafely/hypertension/2020-04-28/ | Ever diagnosis or record prior to study start date |

**Quantitative bias analysis for living with children and COVID study**

We used literature estimates to specify a plausible range of values for the association between COVID-19 and high-risk occupation.

In data from the Labour Force Survey 31% of UK working-age adults with children were key workers, whereas 27% of those without children were key workers. Key workers ^16^ are workers who are deemed by the UK government to be essential during the COVID-19 outbreak. This grouping includes health care workers, social care workers, and others in frontline positions (i.e. police), who, as such, are likely at higher risk of COVID-19 infection.

Based on this literature estimate we specified a range of values for the plausible prevalence of high-risk occupation among those with and without children: 0.25, 0.3, and 0.35.

We also specified a range of plausible values for the association between high-risk occupation and COVID-19 infection.

Among key workers, it might be anticipated that healthcare workers are at highest risk. As such we used the risk ratio between health care worker occupation and COVID-19 seroprevalence in a published Danish study (1.33, 95% CI 1.12-1.58) to specify a plausible worst case range of plausible values for the association between high-risk occupation and COVID-19 infection: 1.3, 1.6, and 2.^17^

We assume the effect of high-risk occupation on COVID-19 outcomes is mediated solely by risk of COVID-19 infection. In UK Office for National Statistics data there was no difference in COVID-19 mortality between health care workers and the working-age population.^18^
